# Modeling and Short-Term Forecasts of Indicators for COVID-19 Outbreak in 25 Countries at the end of March

**DOI:** 10.1101/2020.04.26.20080754

**Authors:** Handan Ankaralı, Nadire Erarslan, Özge Pasin

## Abstract

**Background:** The coronavirus, which originated in Wuhan, causing the disease called COVID-19, spread more than 200 countries and continents end of the March. There is a lot of data since the virus started. However, these data will be explanatory when accurate analyzes are made and will allow future predictions to be made. In this study, it was aimed to model the outbreak with different time series models and also predict the indicators.

**Methods:** The data was collected from 25 countries which have different process at least 20 days. ARIMA(p,d,q), Simple Exponential Smoothing, Holt’s Two Parameter, Brown’s Double Exponential Smoothing Models were used. The prediction and forecasting values were obtained for the countries. Trends and seasonal effects were also evaluated.

**Results:** China has almost under control according to forecasting. The cumulative death prevalence in Italy and Spain will be the highest, followed by the Netherlands, France, England, China, Denmark, Belgium, Brazil and Sweden respectively as of the first week of April. The highest daily case prevalence was observed in Belgium, America, Canada, Poland, Ireland, Netherlands, France and Israel between 10% and 12%.The lowest rate was observed in China and South Korea. Turkey was one of the leading countries in terms of ranking these criteria. The prevalence of the new case and the recovered were higher in Spain than Italy.

**Conclusions:** More accurate predictions for the future can be obtained using time series models with a wide range of data from different countries by modelling real time and retrospective data.

## INTRODUCTION

Coronavirus disease (COVID-19, SARS-CoV-2), which started in Wuhan, China in December 2019, has been recognized as a global threat and it turned into a pandemic that threatened two hundred countries at the end of March. Although there are differences in various indicators such as cumulative case and cumulative death among countries, the outbreak can be estimated with various mathematical models. For this purpose, various models such as logistics model, Bertalanffy model, Gompertz model^1^, Cubic Curve^2^, Exponential Curve, ARIMA models^3^, phenomenological models^4^, optimization algorithms^5^ have been examined before. But the estimates have been made for several countries such as China, Japan and USA, whose data are relatively much more^1–9^. In many countries, the total number of cases reached very high at the end of March, and the outbreak period has exceeded one month. Under these conditions, modeling the data collected from many countries to define the outbreak process and the characteristics of the outbreak, to take precautions and to create projection for the countries where the outbreak has begun or will began is of great importance.

In this study, it was aimed to model the outbreak for six indicators from twenty five countries with different starting dates and processing at least twenty days of data using the end of March data. For this purpose, ARIMA (p, d, q), Simple Exponential Smoothing Model, Holt’s Two Parameter Model, Brown’s Double Exponential Smoothing Models were used. Then prediction and forecasting values were presented for the countries.

## MATERIALS AND METHODS

### Data - Indicators and Models

In the study, twenty five countries were selected which cumulative cases exceeding 1000 among the 160 countries have exposed to the COVID-19 outbreak before March 15 and data for six indicators were modeled. The countries selected for modeling were divided into periods at certain intervals according to the start times of the outbreak and was presented in Table 1 as a summary. The number of countries struggling with the outbreak increased to 200 at the end of March but data from 40 countries that did not contain sufficient data for modeling and started to struggle the outbreak after March 15 were excluded from the study. Since at least one of the countries which was announcing the first case at different periods, was selected for modeling, the results to be obtained from these countries will create a prediction for other countries under similar conditions and for countries that have just started the outbreak.

**Table 1.** The starting periods of the outbreak in the selected countries for modelling

| The Reported First Case(s) | Total Countries | Selected Countries | Number of selected Countries |
| --- | --- | --- | --- |
| Dec 31 - Jan 15 | 3 | China | 1 |
| Jan 16 - Jan 31 | 24 | Italy, Spain, South Korea, Germany, France, USA, UK, Canada, Sweeden, Australia, Malaysia | 11 |
| Feb 1 - Feb 15 | 2 | Belgium | 1 |
| Feb 16 - Feb 20 | 3 | Iran, Israel | 2 |
| Feb 21 – Feb 29 | 37 | Switzerland, Netherlands, Austria, Norway, Brazil, Denmark, Ireland | 7 |
| March 1 - March 7 | 39 | Portugal, Poland | 2 |
| March 8 - March 15 | 52 | Turkey | 1 |

The data were obtained from the internet sources (WHO, Worldometers and Wikipedia). The data of the indicators described below were modeled.

- Cumulative Cases
- Cumulative Deaths
- Daily Cases
- Daily Deaths
- Cumulative Recovered
- Active Cases

### Time Series Models used in the Study

*ARIMA (p,d,q):* ARIMA time series are non-stationary time series, AR, MA and ARMA processes are stationary series. Generally, non-stationary processes are encountered in applications. Because, in practice, data generally varies due to seasonal, irregular movements or incidental reasons. The process becomes stationary when the non-stationary ARIMA time series is taken from the appropriate degree. ARIMA processes are expressed in ARIMA (p, d, q) notation. Here, *p* indicates the degree of autoregressive process, *d* is the difference of degrees, and *q* is the degree of the moving averages process. The ARIMA (p, d, q) process includes both AR (p), MA (q) and ARIMA (p, q) processes. The ARIMA process is an integrated autoregressive moving average process. In the ARIMA models, where d = 1, p and q is 0, then the ARIMA (0,1,0) process is obtained, which is defined as the random walk process^10–12^.

When the first difference of the non-stationary *Y_t_* series is taken (d=1), the 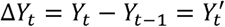 equation is obtained. If the series is not still stationary, a second difference is taken. 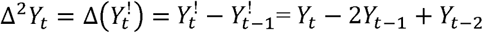 is obtained. In this case, d = 2. Therefore, the non-stationary series is made stationary and shows the ARMA process. The model takes the form of ARIMA (p, d, q). In the method, trial method is used to determine the appropriate *p, d* and *q* values. Therefore, this method has the disadvantage, but it gives very good results in short term estimations. While d value is determined as the number of differences in the series, *p* and *q* values are obtained by examining partial autocorrelation and autocorrelation functions, respectively. The least squares method or nonlinear methods are used in parameter estimates. Assessment of goodness of fit is investigated by AIC and SIC criteria. In addition, compliance with the white noise process is checked for model suitability. The model must comply with the white noise process^10–12^.

*Simple Exponential Smoothing Model:* The advantage of the model is that it can be applied in series with stochastic trends and the estimates can be updated by evaluating the latest changes in the data. The updating is done by giving different weights to the historical data. In cases where periodic and irregular fluctuations are very large, when there is no trend and seasonality, the use of exponential smoothing methods enables more accurate estimations. In the exponential smoothing method, a smoothing coefficient is used and it gives decreasing values according to the distances from today’s period^13^.

According to the seasonal component, the model is *S_t_* = *S_t_*_−1_ + *a*(*Y_t_*_−1_ − *S_t_*_−1_). In the model is the observation value at time *t* and the *S_t_* is the smoothed observation value at time t. The forecast error is equal to *Y_t_*_−1_ − *S_t_*_−1_. The smoothing parameter *a* takes values between 0 and 1. Generally 0.05≤ *a ≤* 0.3 range is used. For optimal selecting smoothing parameters, researchers can perform different range of *a* values. The *a*, which gives the lowest average standard error value, will be the best choice^13^.

The simple exponential smoothing method is only suitable for time series that move around an average. In the moving averages method, equal weight is given to the period values by experiment while different weights are given in the simple exponential smoothing method and these values are becoming exponential shape. In the simple exponential smoothing method, higher values are given to recently obtained values in weighting. The simple exponential smoothing method is often used to estimate short-term predictions^13^.

*Holt’s Two Parameter Model:* The Holt’s two parameter model is used when there is a trend in the data. But in the data there is no seasonality. In the method, while each forecast is obtained, the previous forecast is updated and new forecast values are obtained. Since the method also takes into account the trend in the data, it is slightly more complicated than moving averages and simple exponential smoothing methods^14^.

Holt’s two parameter model is created with the help of the following equations.

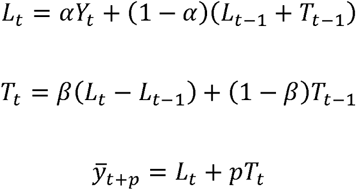

In the above functions, p is the number of periods predicted, while the value of *L_t_* is the new smoothed value, while *T_t_* is the trend estimate value and *Y_t_* is the actual value in period t. The 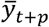 is the post period prediction value^14^.

Method uses trend estimates, so in the model there is two coefficients. α is the smoothing coefficient and the β is the coefficient used for trend estimation. These coefficients ranges between 0 and 1. The alpha and beta coefficients that make the total error square minimum are chosen as the most appropriate parameter value.

*Brown’s Double Exponential Smoothing Model:* This model was obtained as a result of the development of the simple exponential smoothing method. While the series contains trends, it does not include seasonality. Equations of the method is given in the below^15^.

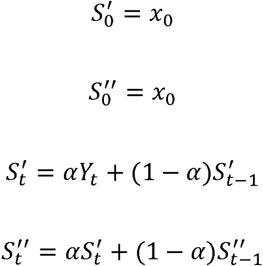

*S′* is the smoothed series and *S″* is the double smoothed series. They obtained by simple exponential smoothing model. For *Y_t_*_+_*_k_* forecasting, the equation is *F_t_*_+_*_k_* = *L_t_* + *kT_t_*.

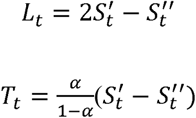

Here *L_t_* is the predicted level in time t, while *T_t_* is the predicted trend in time. If there is an increase and decrease in the series in the data set, Brown’s model gives better results. In the method, two different smoothed series are used, which are located in two different centers. Brown’s method tries to create a linear equation. Estimates include single and double smoothed constants. Therefore, the method takes its name from this feature^15^.

### Statistical Analysis

In modeling, cumulative cases, cumulative deaths, daily cases, daily deaths, cumulative recovered and active cases, which are indicators of the COVID-19 outbreak, were used. The models with the most successful results were selected and 10-day forecasts after the modelling were calculated. In the modelling process, autocorrelation values were calculated in the data and seasonality and trend of the data were examined. The largest R-square value and the smallest RMSE and Normalized BIC values were used for the model performance.

## RESULTS

The results of the China is very important for many countries because the COVID-19 outbreak first began in China. In addition, China has caused the outbreak to spread to many countries and has been fighting this epidemic for 3 months. Also, as of the end of March, it is the fifth country with the highest number of cases and total deaths in the world When the modeling results of China data were investigated considering six different indicators, It was seen that the first four indicators were modeled with high success, and sufficient success was reached in the modeling of cumulative recovered (Table 2). However, there was no suitable time series model that successfully models active cases. (R-squared < 0,50).

**Table 2.** Times series models for China

| Country |  | Model Type | R-squared | RMSE | Normalized BIC |
| --- | --- | --- | --- | --- | --- |
| China | Cumulative Cases | ARIMA(0,2,1) | 1,000 | 253,049 | 11,130 |
|  | Cumulative Deaths | Brown | 1,000 | 16,034 | 5,612 |
|  | Daily Cases | Simple | ,997 | 1660,299 | 14,894 |
|  | Daily Deaths | Simple | ,992 | 1681,084 | 14,919 |
|  | Cumulative Recovered | Brown | ,865 | 16,088 | 5,619 |

The 10-day forecast results after the model building date for the indicators modeled with high success were given in Table 3.

When the table was evaluated, it was observed that the number of cases did not change much except recovered cases in the first half of April in China.

**Table 3.** Forecasting for 10 days in China

| Date | Cumulative Cases | Cumulative Deaths | Daily Cases | Daily Deaths | Cumulative Recovered |
| --- | --- | --- | --- | --- | --- |
| 29.03.20 | 81491 | 3305 | 52 | 5 | 75910 |
| 30.03.20 | 81542 | 3309 | 52 | 5 | 76372 |
| 31.03.20 | 81594 | 3314 | 52 | 5 | 76834 |
| 01.04.20 | 81645 | 3318 | 52 | 5 | 77297 |
| 02.04.20 | 81697 | 3323 | 52 | 5 | 77759 |
| 03.04.20 | 81749 | 3327 | 52 | 5 | 78221 |
| 04.04.20 | 81800 | 3332 | 52 | 5 | 78684 |
| 05.04.20 | 81852 | 3336 | 52 | 5 | 79146 |
| 06.04.20 | 81903 | 3341 | 52 | 5 | 79609 |
| 07.04.20 | 81955 | 3345 | 52 | 5 | 80071 |

Estimates of different indicator data obtained from China are shown in Figure 1. From the graphs, it was seen that there was no significant change in the number of cumulative cases and the number of cumulative deaths as of the end of March. But the number of cumulative recovered was seen to increase. Daily cases and daily death were close to the minimum level.

**Figure 1.**
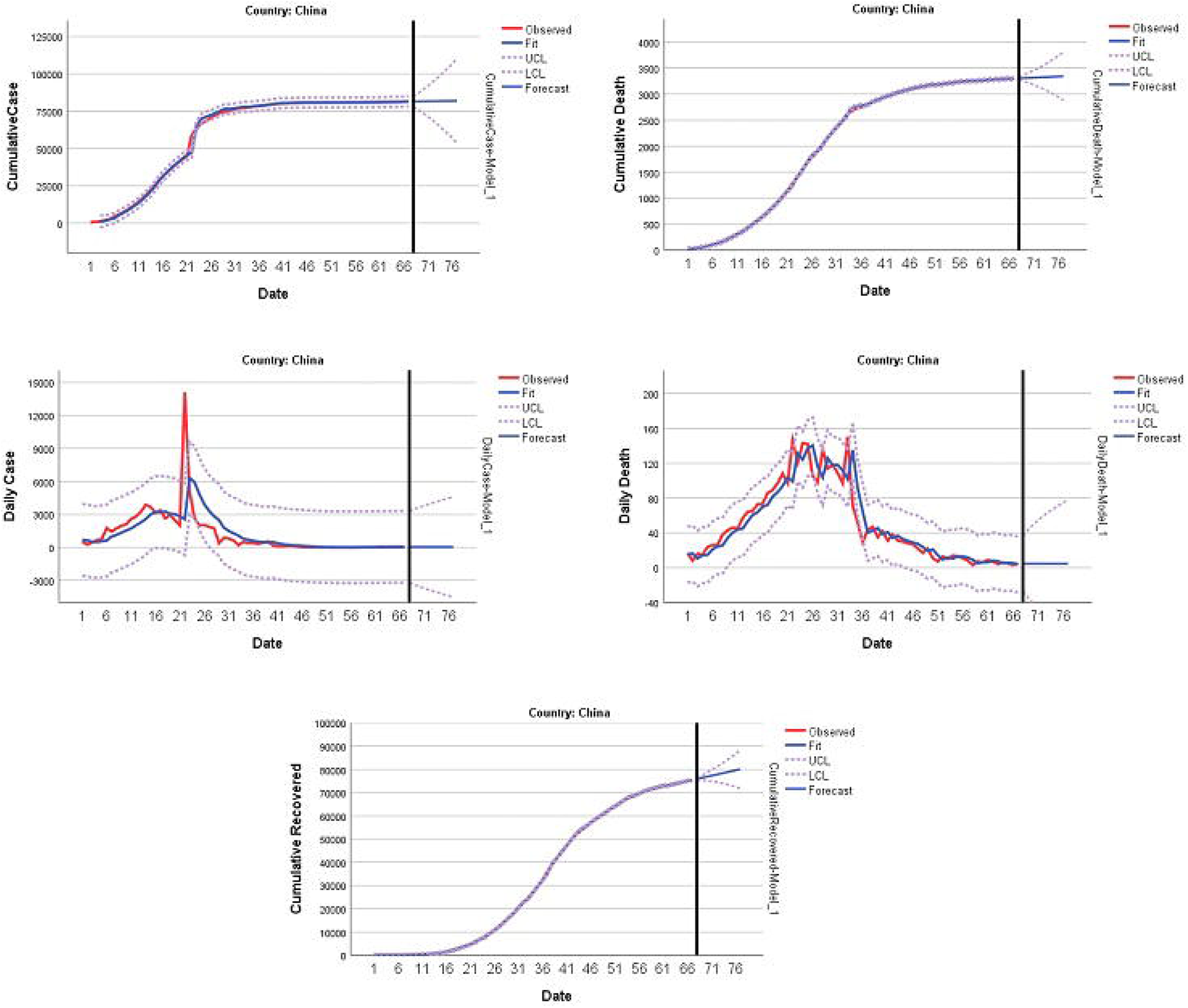
Predicting and forecasting of indicators for China

In 11 out of 24 countries that detected the first Coronavirus cases in the period of 16-31 January, the total number of cases was over 1000. So the data of the indicators used in the prediction of the outbreak process were modeled and the results are presented in Table 4 and Table 5. It was seen that the activated cases can be modeled successfully in 6 out of 11 countries, and there was no time series model that produces a successful forecast for other countries (Table 4, R-squared < 0,50). In addition, success in modeling the number of cumulative recovered numbers in several countries was between 70% and 85%, while the prediction success of models for other indicators was calculated around 99%.

**Table 4.**
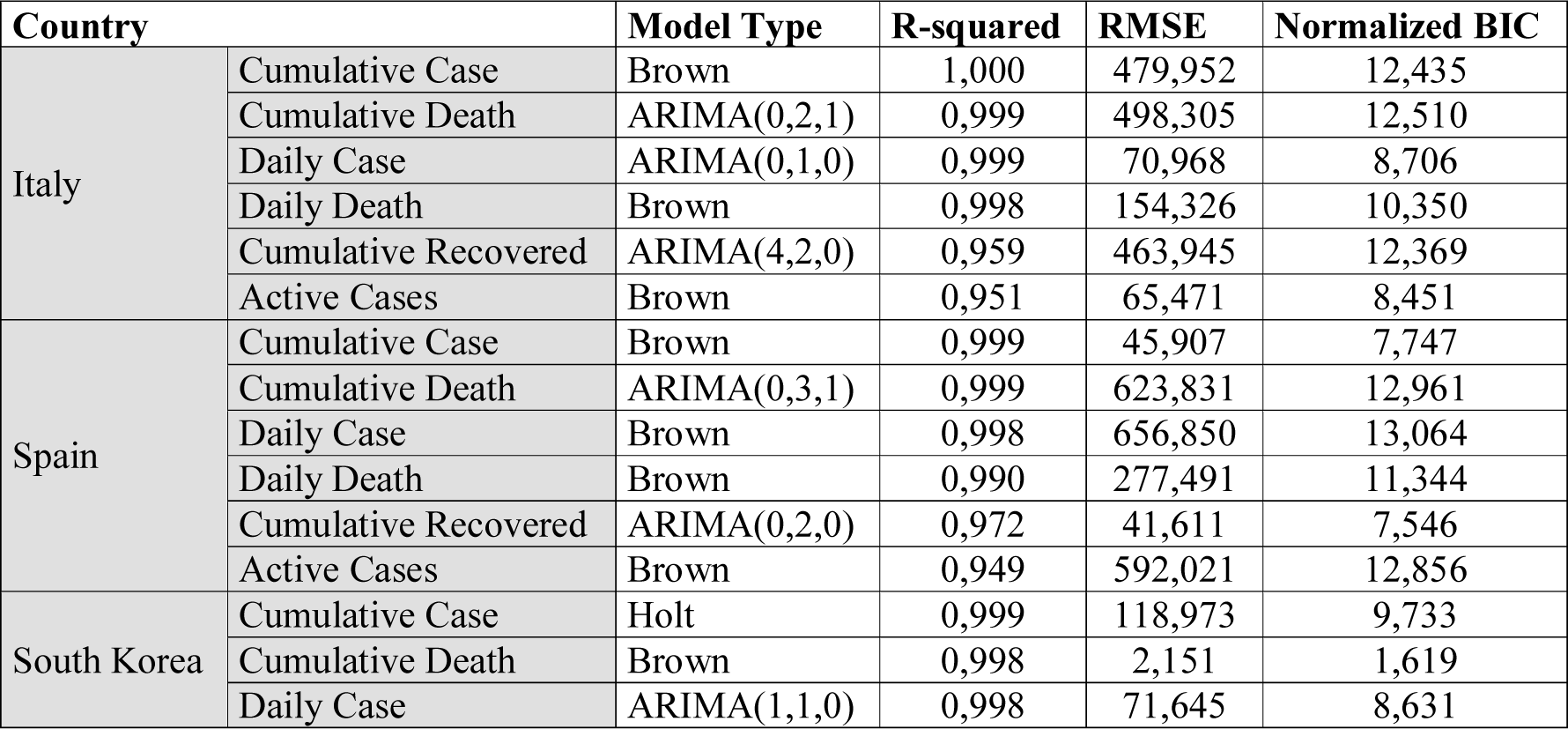
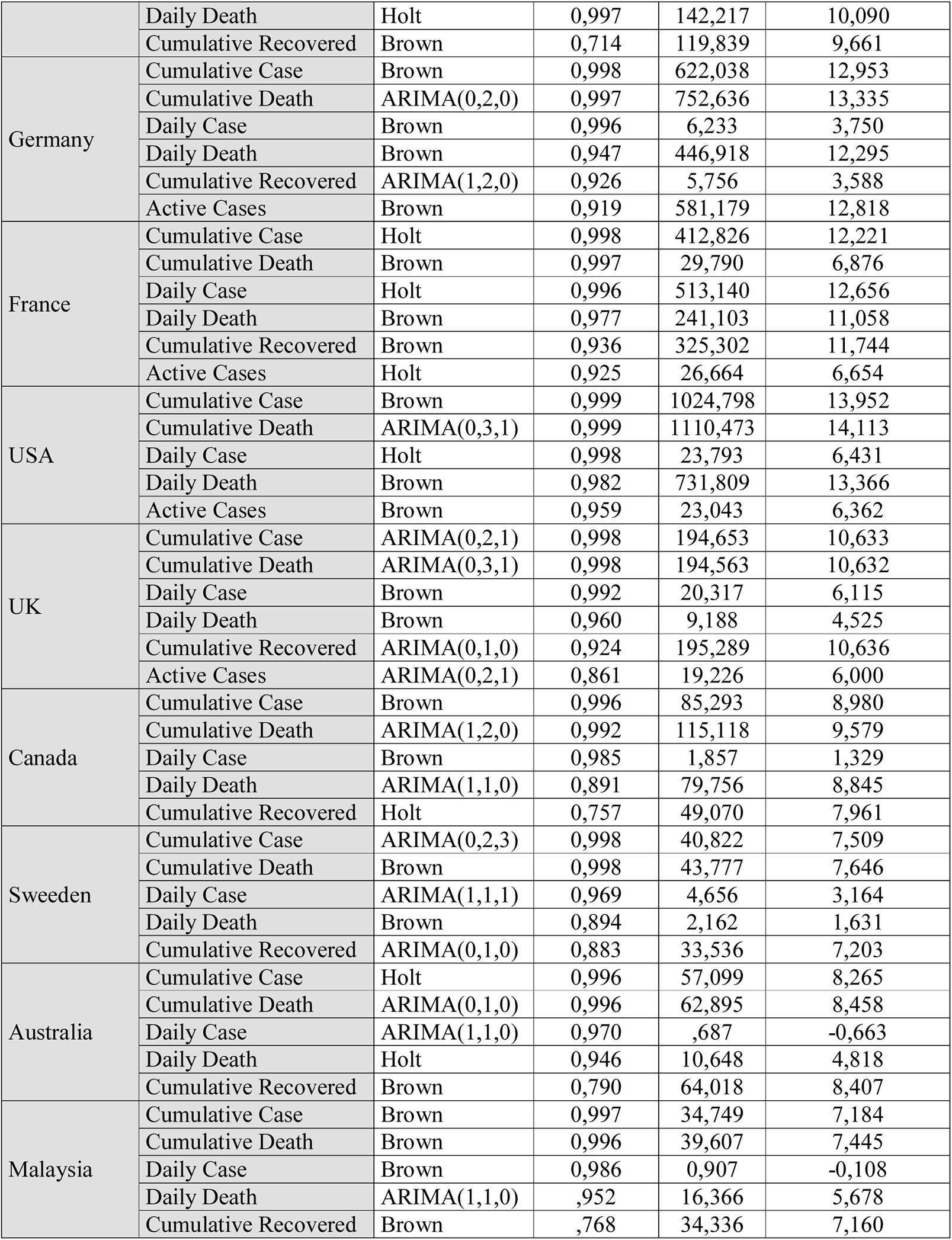
Times series models for the countries that announced the first case in the period of 16-31 Jan

**Table 5.** Forecasting for 10 days in Italy

| Date | Cumulative Cases | Cumulative Deaths | Daily Cases | Daily Deaths | Cumulative Recovered | Cumulative Active Cases |
| --- | --- | --- | --- | --- | --- | --- |
| 29.03.2020 | 98446 | 10930 | 6116 | 928 | 13419 | 73894 |
| 30.03.2020 | 104420 | 11858 | 6258 | 970 | 14385 | 77712 |
| 31.03.2020 | 110394 | 12808 | 6401 | 1012 | 15349 | 81530 |
| 01.04.2020 | 116368 | 13781 | 6543 | 1054 | 16803 | 85348 |
| 02.04.2020 | 122342 | 14775 | 6685 | 1097 | 17665 | 89166 |
| 03.04.2020 | 128316 | 15791 | 6827 | 1139 | 18738 | 92984 |
| 04.04.2020 | 134290 | 16829 | 6970 | 1181 | 19896 | 96802 |
| 05.04.2020 | 140264 | 17888 | 7112 | 1223 | 21167 | 100620 |
| 06.04.2020 | 146238 | 18970 | 7254 | 1266 | 21993 | 104438 |
| 07.04.2020 | 152212 | 20073 | 7396 | 1308 | 23228 | 108256 |

Italy was the second country with the highest number of cases in the world and the country with the highest number of deaths as of the end of March. In general, it was observed that the increase in indicator results continues, but the prevalence in the daily cases and daily death was slower than other indicators when Table 5 and Figure 2, which include the results of the forecast for the next 10 days, were examined.

**Figure 2.**
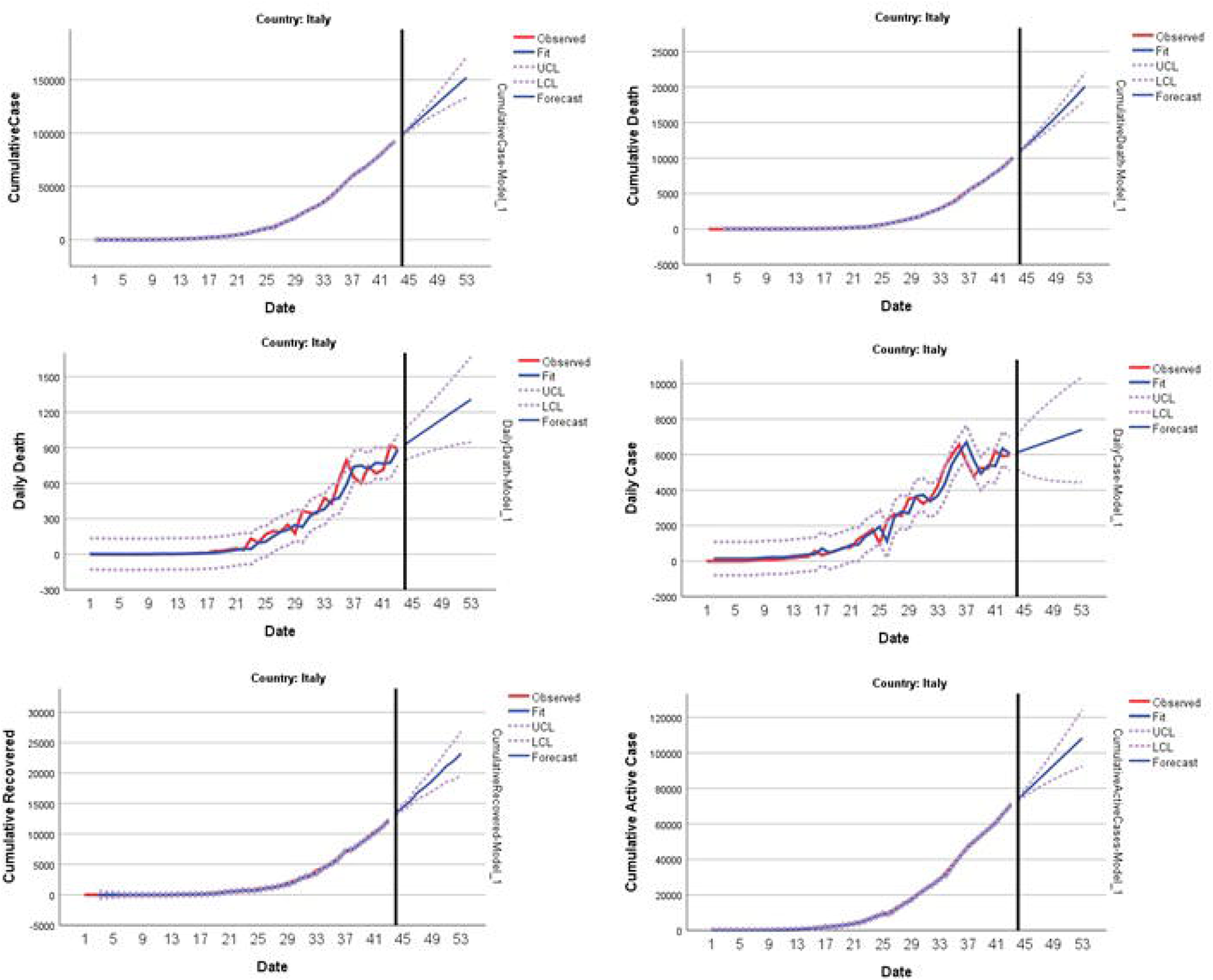
Predicting and forecasting of indicators for Italy

Spain was the 2nd country with the highest total number of deaths and the 4th country with the total number of cases. The forecast values for 10 days after modeling were presented in Table 6 and Figure 3. It can be seen from the graphs that the number of cases and deaths continues to increase rapidly.

**Table 6.** Forecasting for 10 days in Spain

| Date | Cumulative Cases | Cumulative Deaths | Daily Cases | Daily Deaths | Cumulative Recovered | Cumulative Active Cases |
| --- | --- | --- | --- | --- | --- | --- |
| 29.03.2020 | 80763 | 6887 | 8542 | 907 | 15284 | 58975 |
| 30.03.2020 | 88290 | 7852 | 8973 | 970 | 18359 | 62966 |
| 31.03.2020 | 95818 | 8878 | 9403 | 1033 | 21506 | 66956 |
| 01.04.2020 | 103345 | 9965 | 9834 | 1096 | 24727 | 70947 |
| 02.04.2020 | 110873 | 11112 | 10265 | 1159 | 28021 | 74938 |
| 03.04.2020 | 118400 | 12320 | 10695 | 1222 | 31388 | 78928 |
| 04.04.2020 | 125928 | 13589 | 11126 | 1285 | 34829 | 82919 |
| 05.04.2020 | 133455 | 14918 | 11557 | 1348 | 38342 | 86909 |
| 06.04.2020 | 140983 | 16309 | 11987 | 1411 | 41929 | 90900 |
| 07.04.2020 | 148510 | 17759 | 12418 | 1474 | 45589 | 94891 |

**Figure 3.**
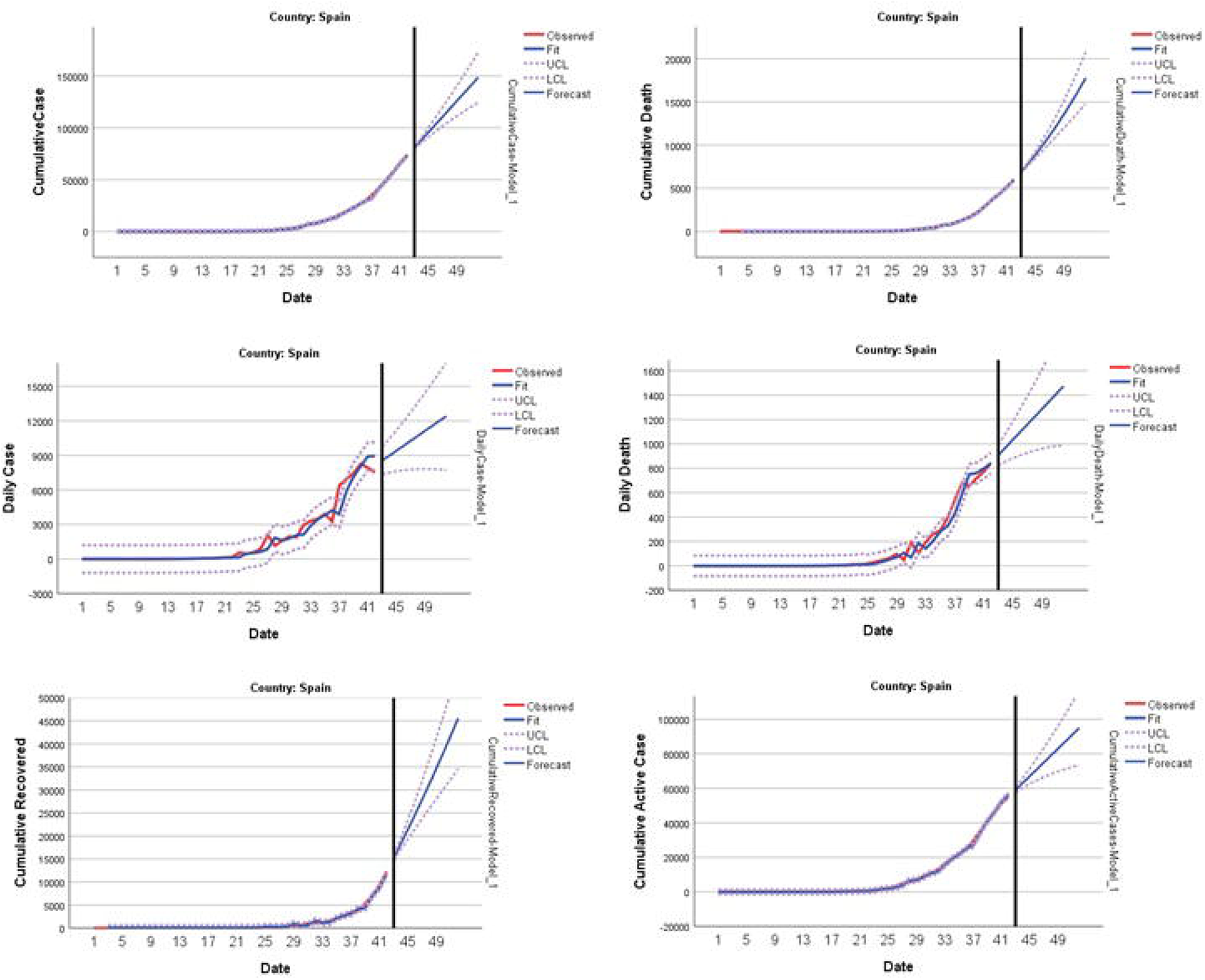
Predicting and forecasting of indicators for Spain

South Korea was a country that has controlled the number of cumulative cases and cumulative deaths since the outbreak. The number of new deaths per day has never exceeded 10. After modeling the obtained data, the 10-day predicted results were given in Table 7 and Figure 4. It was predicted that the changes in the indicators will not change in the first half of April when the table is evaluated.

**Table 7.** Forecasting for 10 days in South Korea

| Date | Cumulative Cases | Cumulative Deaths | Daily Cases | Daily Deaths | Cumulative Recovered |
| --- | --- | --- | --- | --- | --- |
| 29.03.20 | 9603 | 151 | 119 | 6 | 5142 |
| 30.03.20 | 9737 | 157 | 132 | 7 | 5474 |
| 31.03.20 | 9871 | 163 | 126 | 7 | 5805 |
| 01.04.20 | 10005 | 169 | 129 | 7 | 6137 |
| 02.04.20 | 10139 | 175 | 127 | 7 | 6468 |
| 03.04.20 | 10273 | 182 | 128 | 7 | 6800 |
| 04.04.20 | 10408 | 188 | 128 | 7 | 7131 |
| 05.04.20 | 10542 | 194 | 128 | 7 | 7463 |
| 06.04.20 | 10676 | 200 | 128 | 8 | 7794 |
| 07.04.20 | 10810 | 207 | 128 | 8 | 8126 |

**Figure 4.**
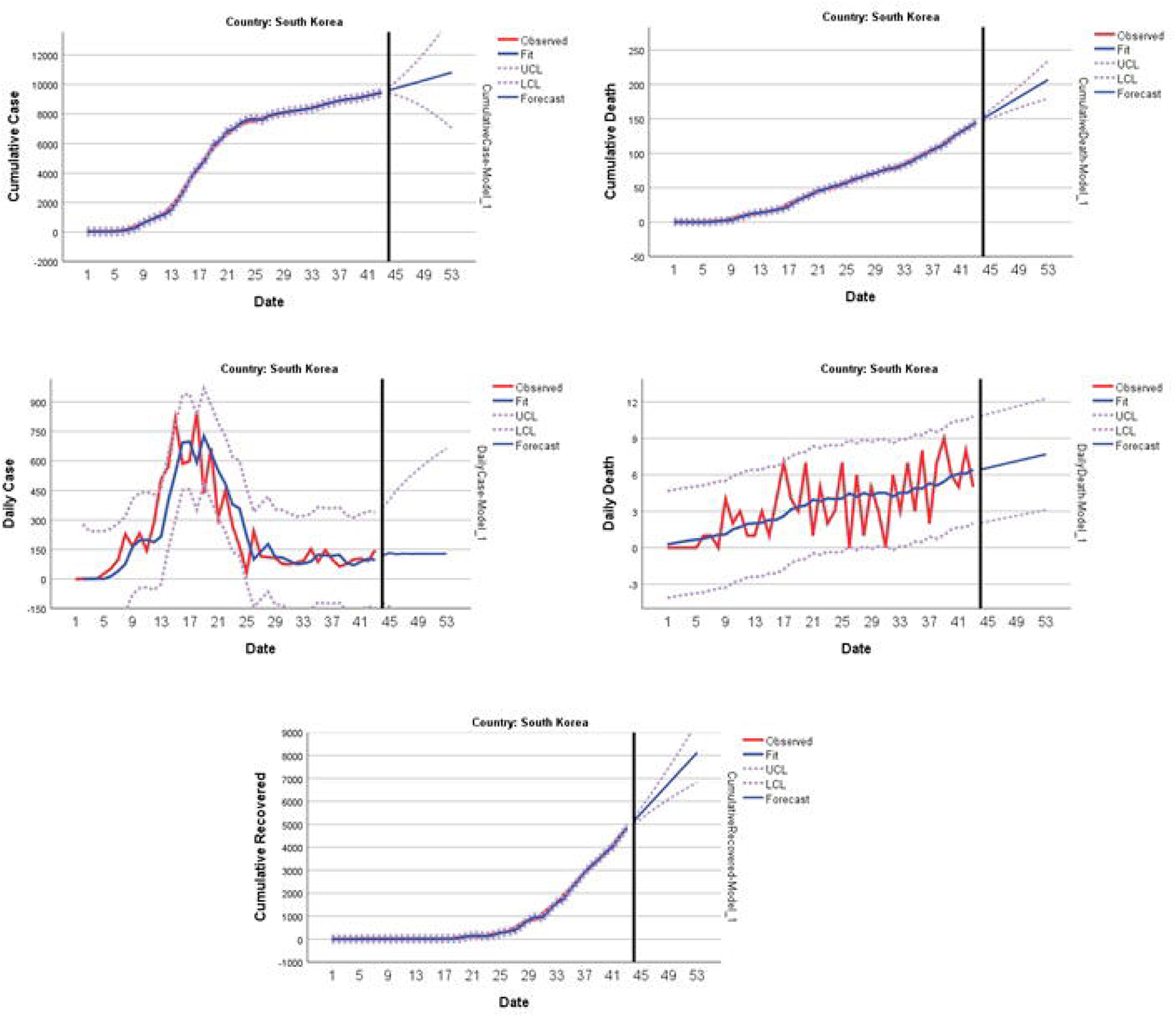
Predicting and forecasting of indicators for South Korea

Germany has been the most successful European country that managed the outbreak for a long time. When the 10-day forecast values given in Table 8 and the forecast of the models in Figure 5 were examined, it was seen that there will be an increase in the indicators similar to the current days and the cumulative death in the cumulative case is very low (1%).

**Table 8.** Forecasting for 10 days in Germany

| Date | Cumulative Cases | Cumulative Deaths | Daily Cases | Daily Deaths | Cumulative Recovered | Cumulative Active Cases |
| --- | --- | --- | --- | --- | --- | --- |
| 29.03.20 | 64525 | 517 | 7476 | 95 | 9664 | 53860 |
| 30.03.20 | 71356 | 603 | 8000 | 106 | 11334 | 58929 |
| 31.03.20 | 78186 | 691 | 8524 | 116 | 12637 | 63999 |
| 01.04.20 | 85016 | 781 | 9047 | 127 | 14219 | 69068 |
| 02.04.20 | 91847 | 873 | 9571 | 137 | 15588 | 74137 |
| 03.04.20 | 98677 | 967 | 10094 | 148 | 17119 | 79207 |
| 04.04.20 | 105507 | 1063 | 10618 | 158 | 18527 | 84276 |
| 05.04.20 | 112338 | 1161 | 11141 | 169 | 20029 | 89346 |
| 06.04.20 | 119168 | 1261 | 11665 | 179 | 21459 | 94415 |
| 07.04.20 | 125998 | 1363 | 12188 | 190 | 22944 | 99485 |

**Figure 5.**
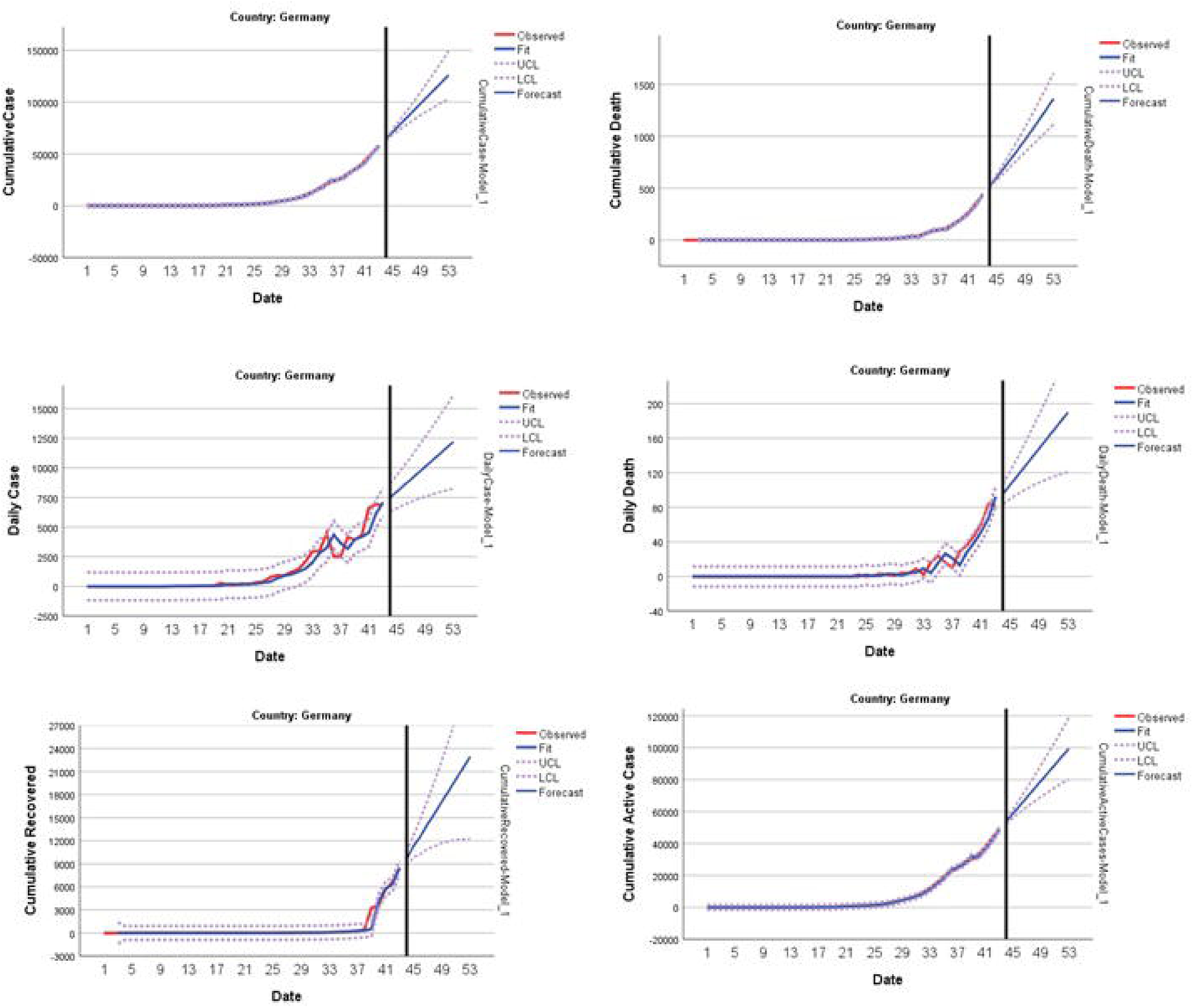
Predicting and forecasting of indicators for Germany

The results for France were summarized in Table 9 and Figure 6. The increase rate in the analyzed indicators continues. In the first half of April, the cumulative recovered prevalence in the cumulative case were expected to be around 14%. The mortality rate in positive cases is expected around 6%.

**Table 9.** Forecasting for 10 days in France

| Date | Cumulative Cases | Cumulative Deaths | Daily Cases | Daily Deaths | Cumulative Recovered | Cumulative Active Cases |
| --- | --- | --- | --- | --- | --- | --- |
| 29.03.20 | 41837 | 2631 | 4773 | 360 | 6390 | 32115 |
| 30.03.20 | 46346 | 2948 | 5245 | 386 | 6935 | 35693 |
| 31.03.20 | 50855 | 3265 | 5717 | 412 | 7479 | 39271 |
| 01.04.20 | 55364 | 3581 | 6189 | 438 | 8024 | 42848 |
| 02.04.20 | 59873 | 3898 | 6661 | 464 | 8569 | 46426 |
| 03.04.20 | 64382 | 4215 | 7133 | 490 | 9113 | 50004 |
| 04.04.20 | 68891 | 4532 | 7605 | 516 | 9658 | 53581 |
| 05.04.20 | 73400 | 4849 | 8077 | 542 | 10202 | 57159 |
| 06.04.20 | 77909 | 5166 | 8549 | 568 | 10747 | 60737 |
| 07.04.20 | 82418 | 5483 | 9021 | 594 | 11292 | 64314 |

**Figure 6.**
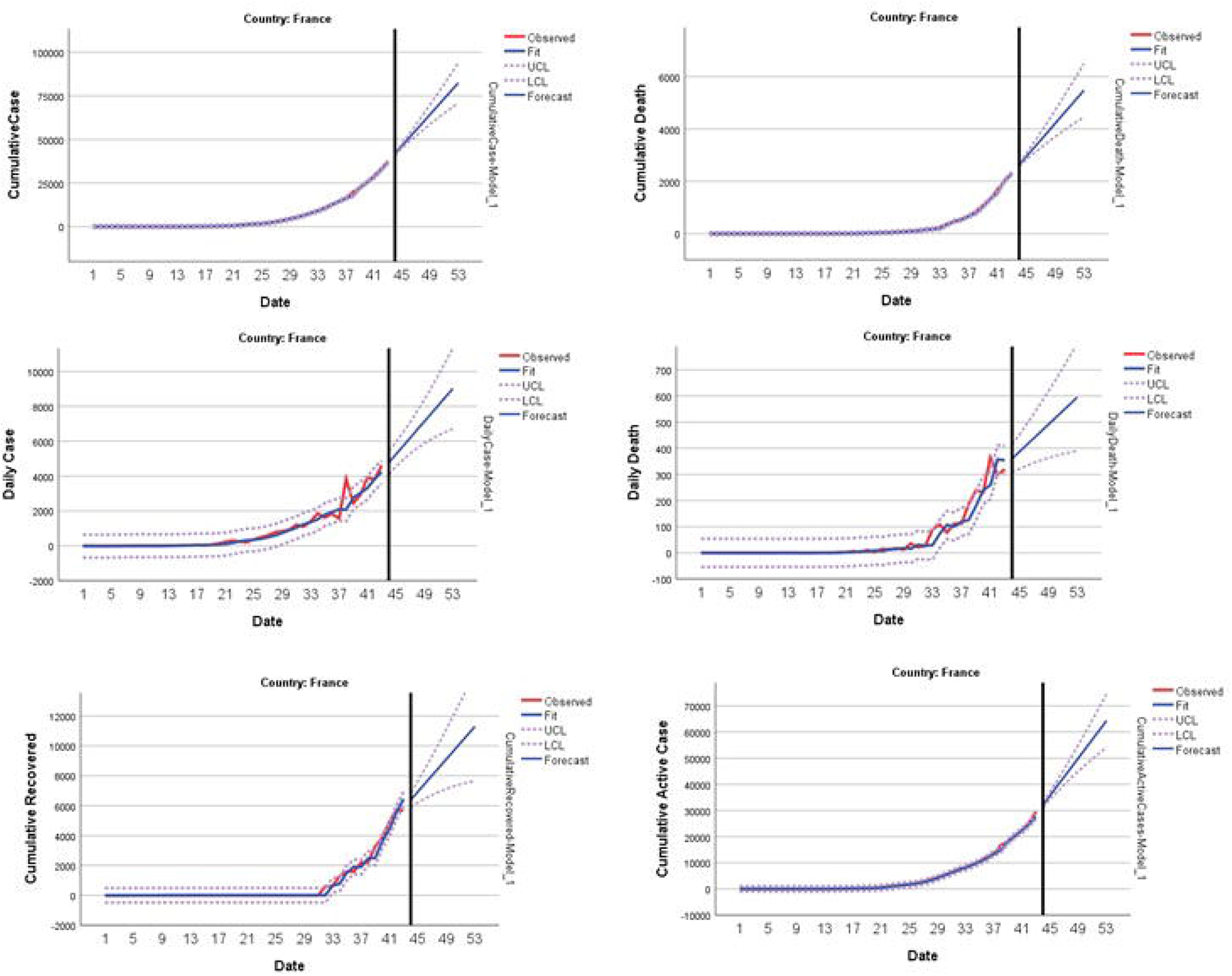
Predicting and forecasting of indicators for France

USA data results were given in Table 10 and Figure 7. The increase rate in the analyzed indicators continues. In the first half of April, the daily case prevalence in cumulative case was expected to be 12% and the cumulative deaths prevalence is around 2%.

**Table 10.** Forecasting for 10 days in USA

| Date | Cumulative Cases | Cumulative Deaths | Daily Cases | Daily Deaths | Cumulative Active Cases |
| --- | --- | --- | --- | --- | --- |
| 29.03.20 | 143030 | 2849 | 22619 | 630 | 136344 |
| 30.03.20 | 162482 | 3579 | 24761 | 739 | 154562 |
| 31.03.20 | 181934 | 4411 | 26903 | 848 | 172780 |
| 01.04.20 | 201386 | 5346 | 29045 | 956 | 190998 |
| 02.04.20 | 220838 | 6384 | 31187 | 1065 | 209216 |
| 03.04.20 | 240290 | 7524 | 33329 | 1174 | 227434 |
| 04.04.20 | 259741 | 8767 | 35471 | 1282 | 245652 |
| 05.04.20 | 279193 | 10112 | 37612 | 1391 | 263870 |
| 06.04.20 | 298645 | 11560 | 39754 | 1500 | 282088 |
| 07.04.20 | 318097 | 13111 | 41896 | 1609 | 300306 |

**Figure 7.**
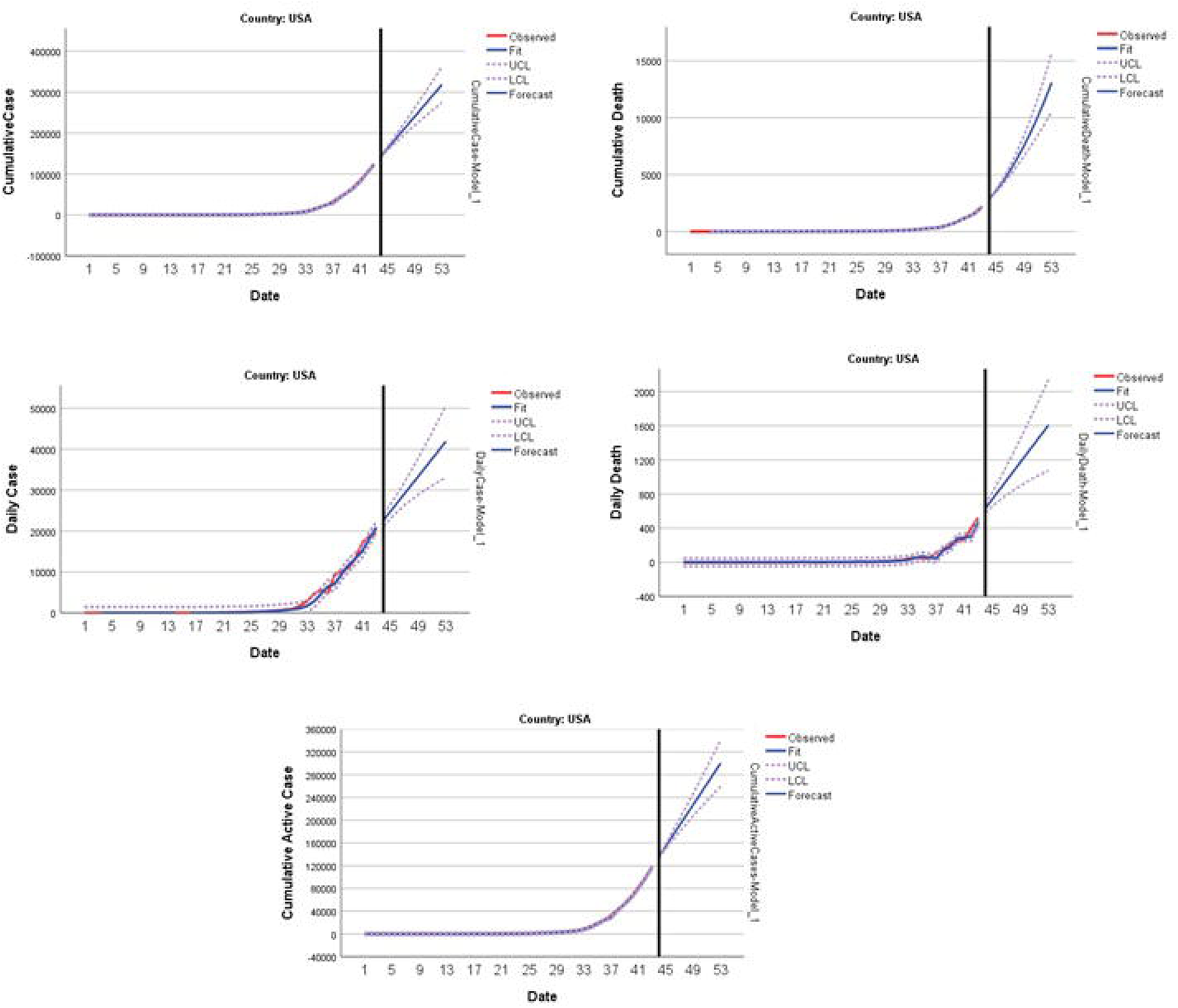
Predicting and forecasting of indicators for USA

The results of UK data were given in Table 11 and Figure 8. The rate of increase in the indicator values examined was similar in the first week of April. In this period, the cumulative recovered prevalence was quite low. The cumulative active case prevalence was found to be quite high (90%).

**Table 11.**
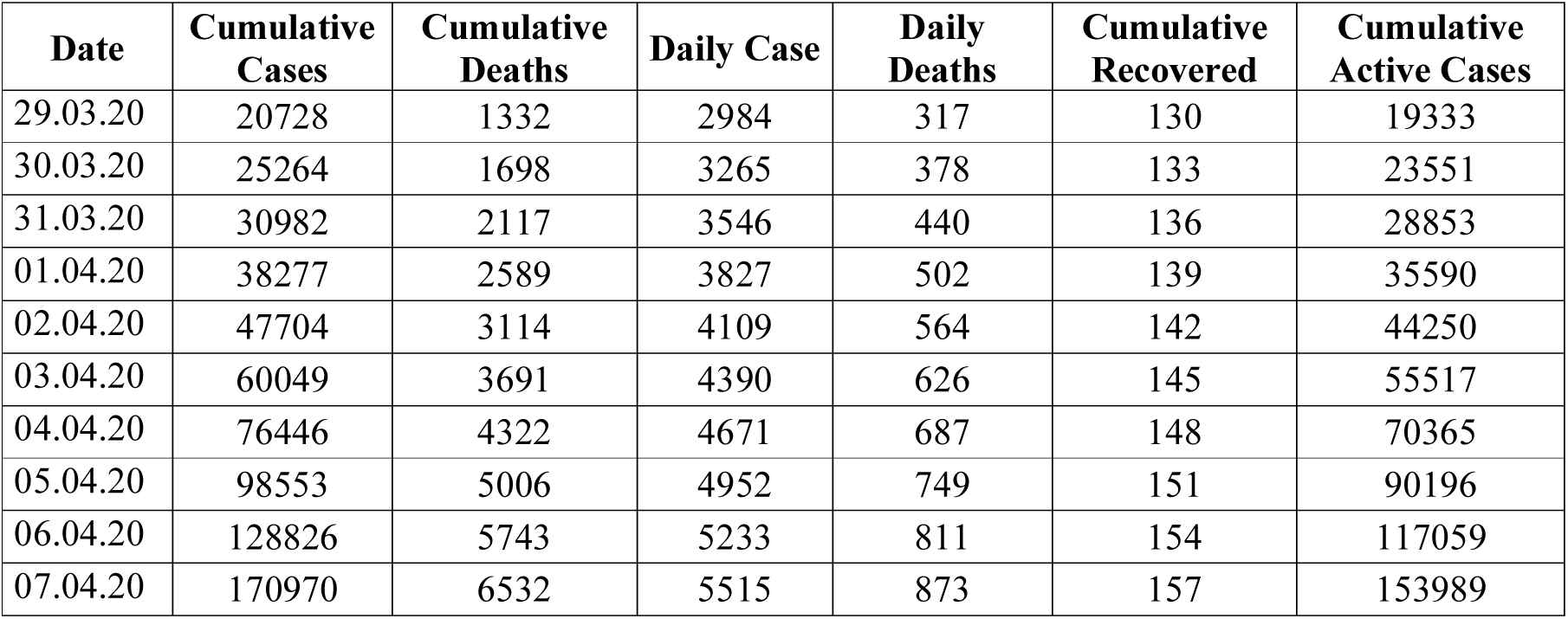
Forecasting for 10 days in UK

**Figure 8.**
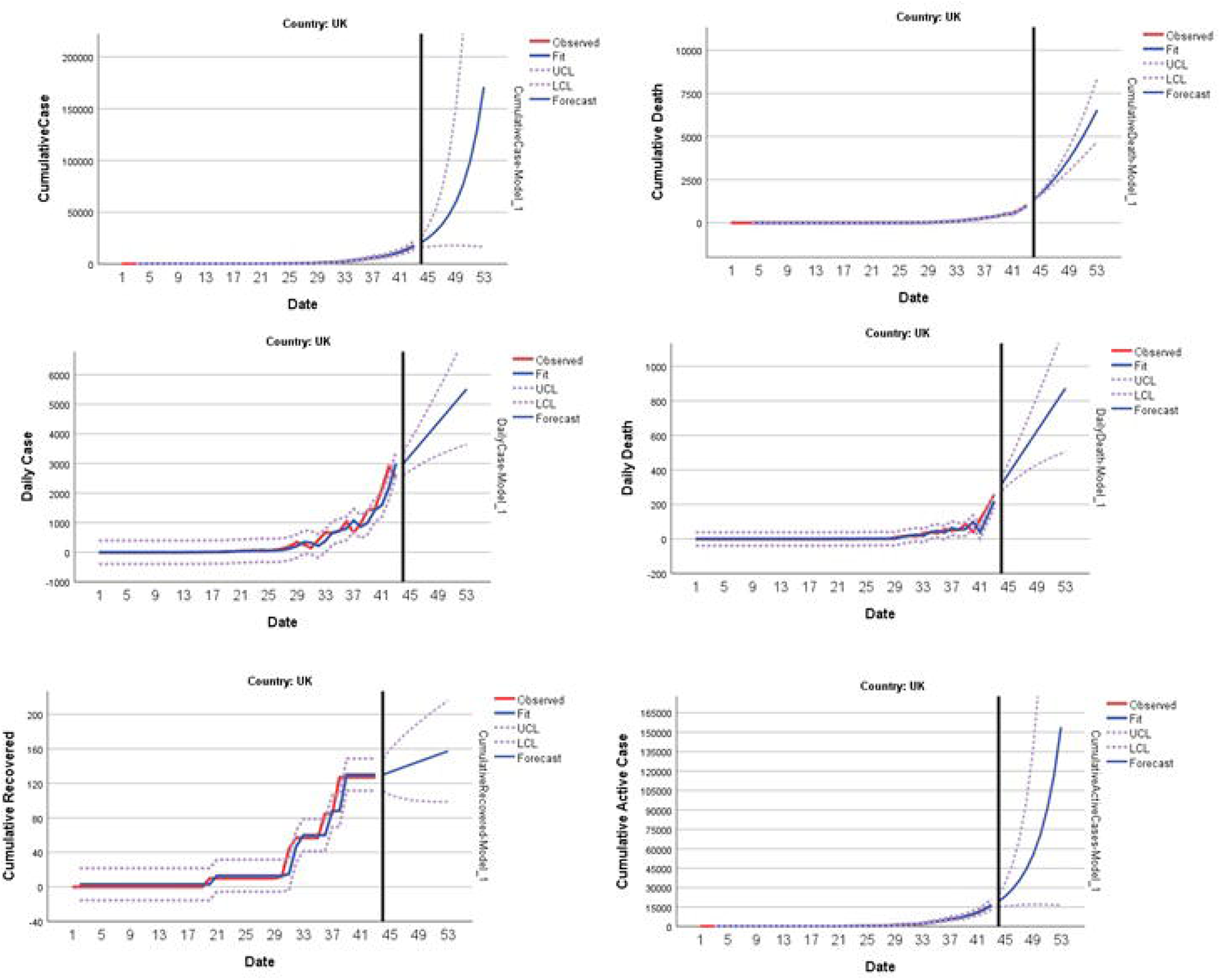
Predicting and forecasting of indicators for UK

When Canada data was evaluated, it is seen that the number of daily deaths is estimated to be quite low (Table 12 and Figure 9). The cumulative recovered prevalence was expected to be around 7% and the cumulative active case prevalence was around 87% in the first week of April.

**Table 12.**
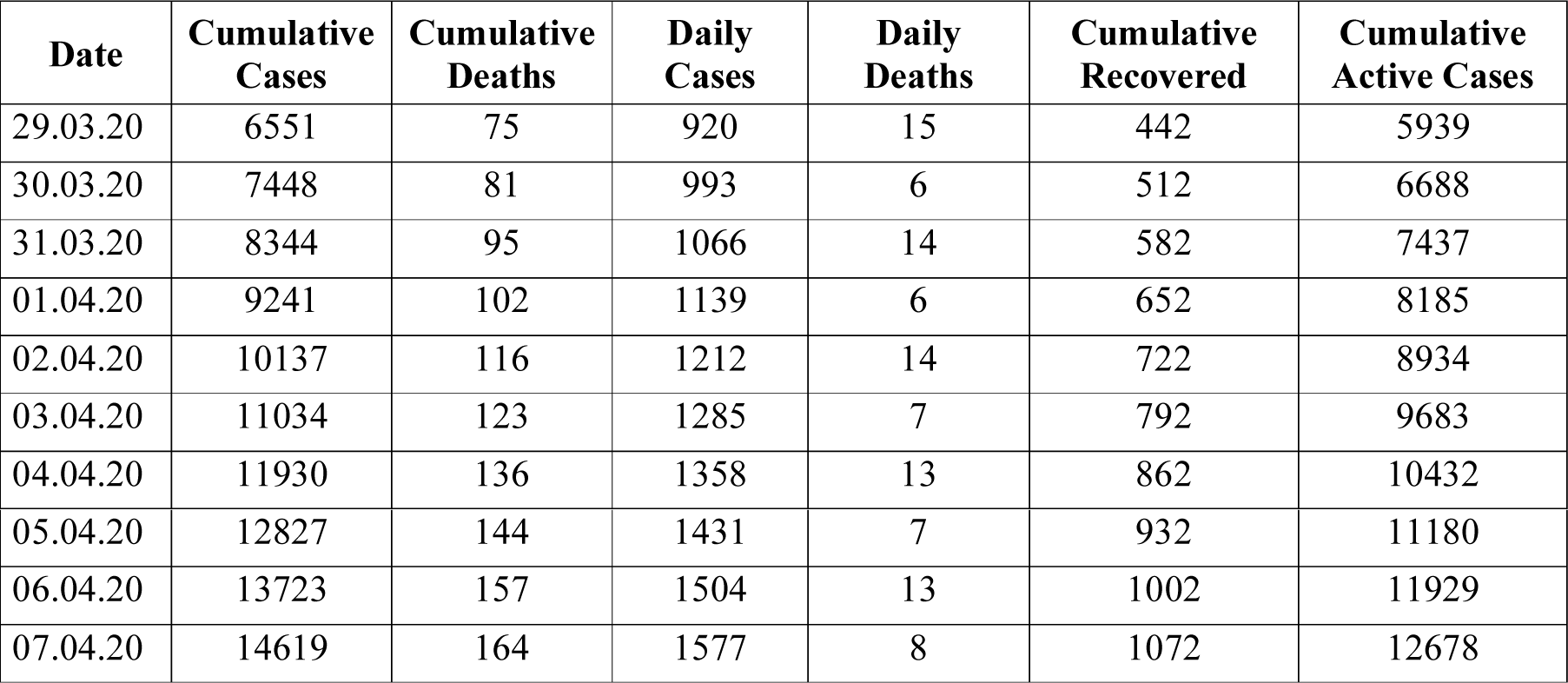
Forecasting for 10 days in Canada

| Date | Cumulative Cases | Cumulative Deaths | Daily Cases | Daily Deaths | Cumulative Recovered | Cumulative Active Cases |
| --- | --- | --- | --- | --- | --- | --- |
| 29.03.20 | 6551 | 75 | 920 | 15 | 442 | 5939 |
| 30.03.20 | 7448 | 81 | 993 | 6 | 512 | 6688 |
| 31.03.20 | 8344 | 95 | 1066 | 14 | 582 | 7437 |
| 01.04.20 | 9241 | 102 | 1139 | 6 | 652 | 8185 |
| 02.04.20 | 10137 | 116 | 1212 | 14 | 722 | 8934 |
| 03.04.20 | 11034 | 123 | 1285 | 7 | 792 | 9683 |
| 04.04.20 | 11930 | 136 | 1358 | 13 | 862 | 10432 |
| 05.04.20 | 12827 | 144 | 1431 | 7 | 932 | 11180 |
| 06.04.20 | 13723 | 157 | 1504 | 13 | 1002 | 11929 |
| 07.04.20 | 14619 | 164 | 1577 | 8 | 1072 | 12678 |

**Figure 9.**
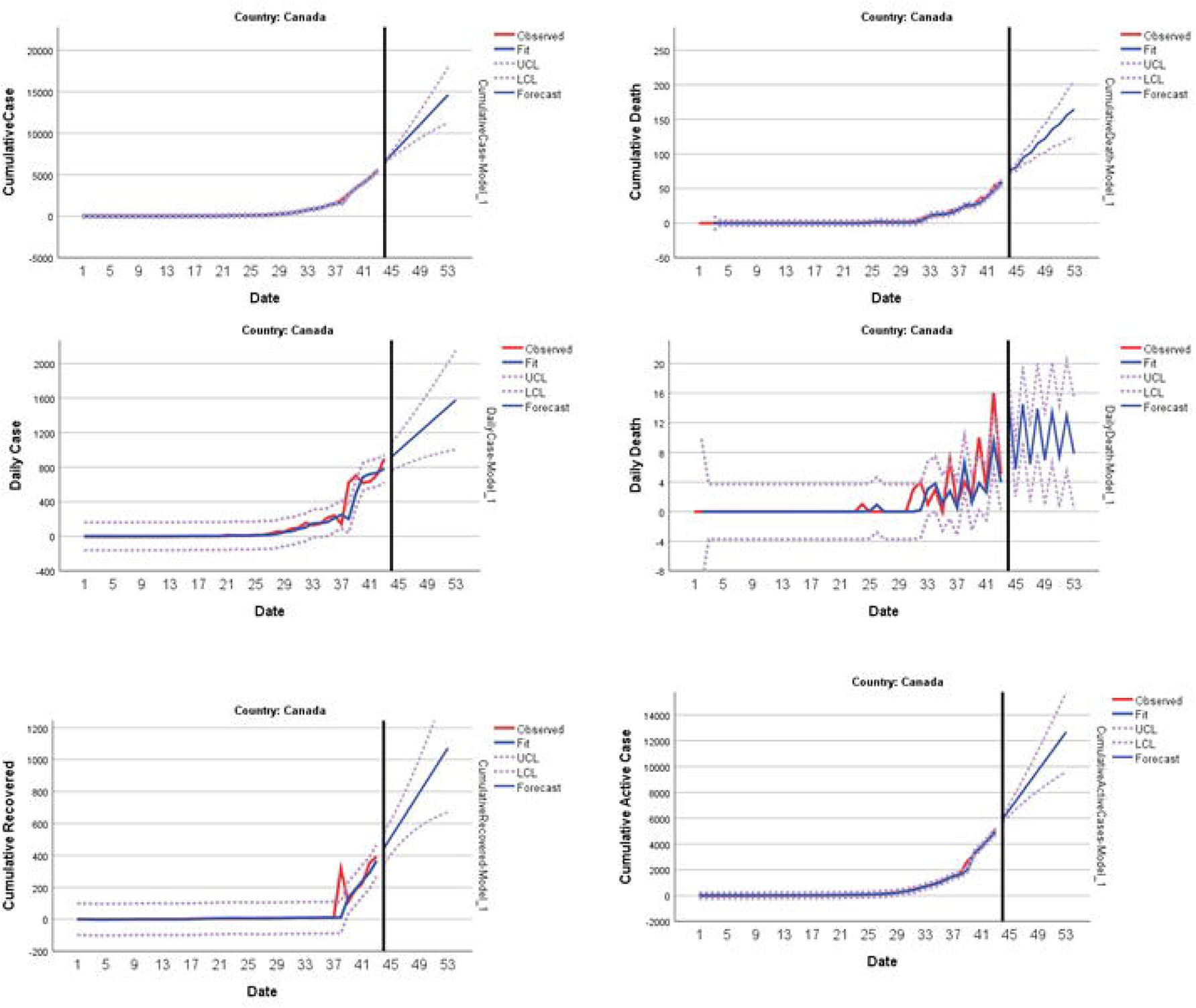
Predicting and forecasting of indicators for Canada

When Sweeden’s data was evaluated, it was predicted that the new daily case prevalence will be around 5% in the first week of April, and the daily mortality and recoveried prevalence will still be at a very low level. In addition, cumulative death prevalence in cumulative case was estimated at 4% levels (Table 13 and Figure 10).

**Table 13.** Forecasting for 10 days in Sweeden

| Date | Cumulative Cases | Cumulative Deaths | Daily Cases | Daily Deaths | Cumulative Recovered |
| --- | --- | --- | --- | --- | --- |
| 29.03.20 | 3822 | 120 | 288 | 14 | 15 |
| 30.03.20 | 4242 | 132 | 377 | 15 | 16 |
| 31.03.20 | 4633 | 144 | 289 | 16 | 16 |
| 01.04.20 | 5042 | 157 | 375 | 17 | 16 |
| 02.04.20 | 5469 | 169 | 290 | 17 | 17 |
| 03.04.20 | 5916 | 182 | 374 | 18 | 17 |
| 04.04.20 | 6383 | 194 | 292 | 19 | 17 |
| 05.04.20 | 6870 | 206 | 373 | 20 | 18 |
| 06.04.20 | 7376 | 219 | 293 | 20 | 18 |
| 07.04.20 | 7904 | 231 | 371 | 21 | 19 |

**Figure 10.**
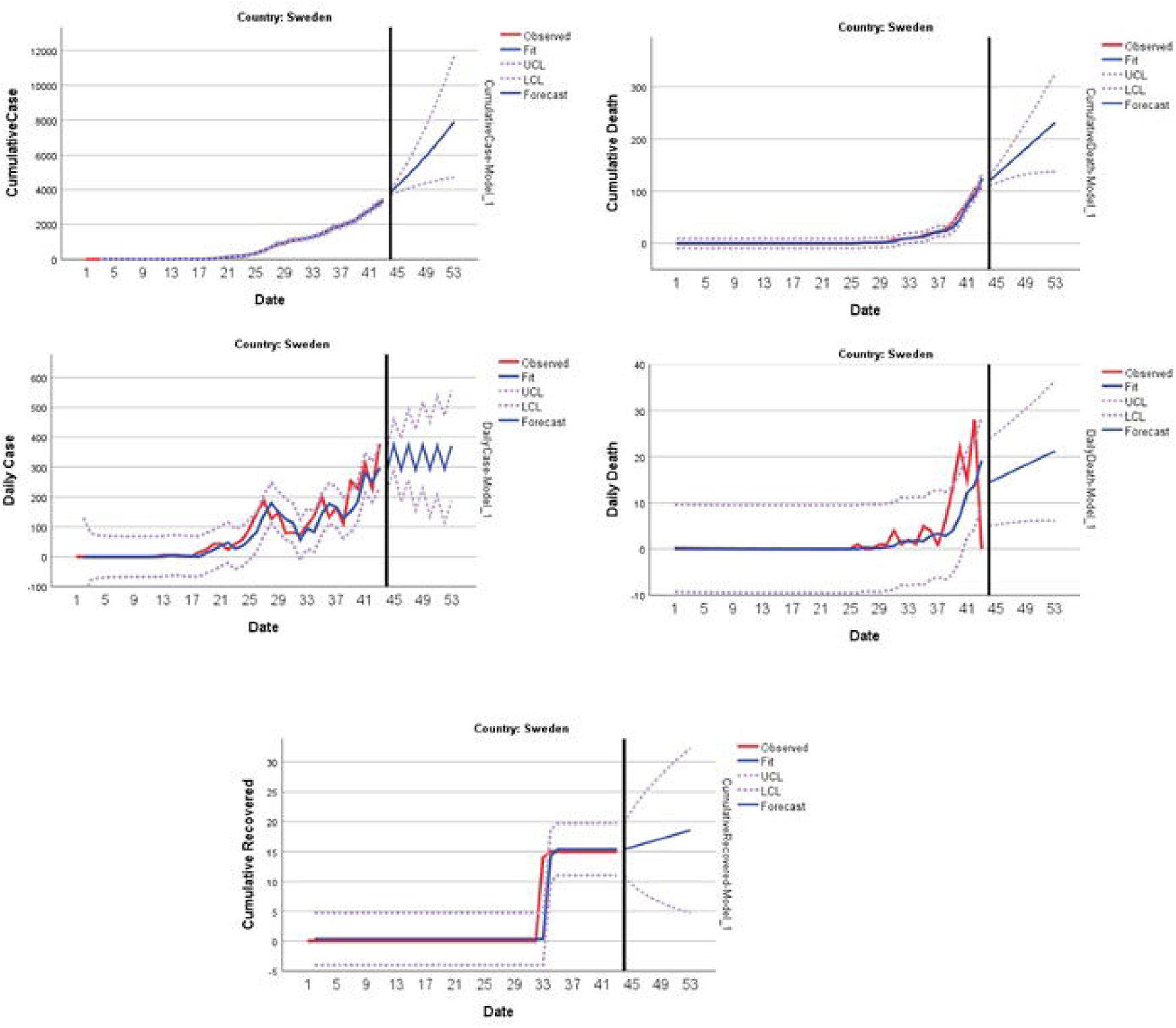
Predicting and forecasting of indicators for Sweden

At the end of the modeling for Australia data, it was predicted that the number of daily deaths will be very low in the first week of April, and the cumulative death prevalence will be observed at a very low level in the cumulative case. In addition, daily new cases and cumulative recovered prevalence were estimated to be around 4% (Table 14 and Figure 11).

**Table 14.** Forecasting for 10 days in Australia

| Date | Cumulative Cases | Cumulative Deaths | Daily Cases | Daily Deaths | Cumulative Recovered |
| --- | --- | --- | --- | --- | --- |
| 29.03.20 | 3934 | 14 | 300 | 1 | 180 |
| 30.03.20 | 4205 | 15 | 274 | 1 | 193 |
| 31.03.20 | 4476 | 15 | 290 | 1 | 207 |
| 01.04.20 | 4747 | 15 | 280 | 1 | 220 |
| 02.04.20 | 5019 | 16 | 286 | 1 | 234 |
| 03.04.20 | 5290 | 16 | 283 | 1 | 247 |
| 04.04.20 | 5561 | 16 | 285 | 1 | 261 |
| 05.04.20 | 5832 | 17 | 283 | 1 | 274 |
| 06.04.20 | 6103 | 17 | 284 | 1 | 288 |
| 07.04.20 | 6374 | 17 | 284 | 1 | 301 |

**Figure 11.**
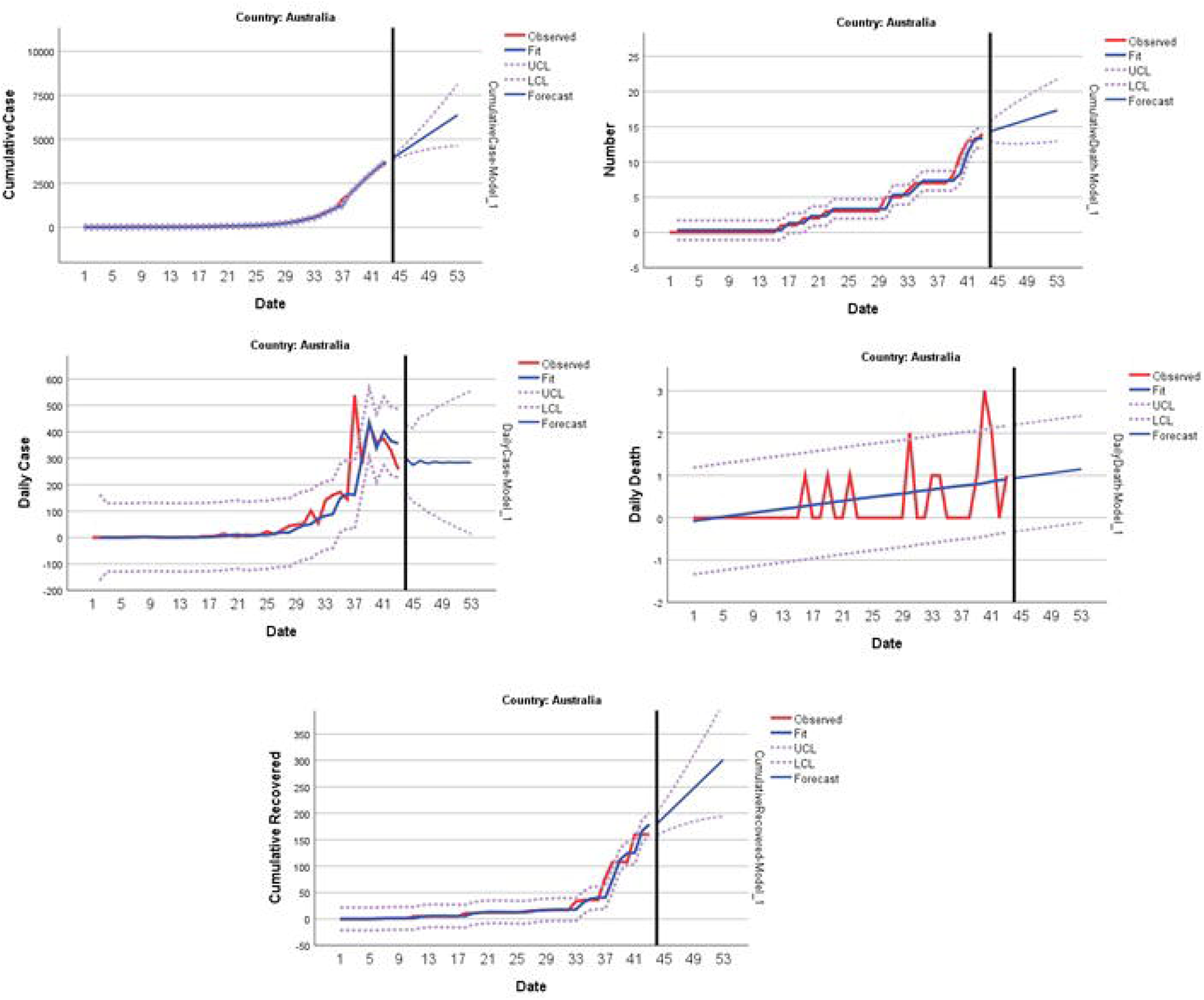
Predicting and forecasting of indicators for Australia

When Malaysia results were analyzed, it was predicted that the number of daily deaths will be very low and the cumulative death prevalence in the cumulative case will be 1% in the first week of April. In addition, the daily case prevalence was estimated to be around 6% and the cumulative recovered prevalence around 15% (Table 15 and Figure 12). These results emphasized that Malaysia has shown successful results in struggle with the outbreak.

**Table 15.** Forecasting for 10 days in Malaysia

| Date | Cumulative Cases | Cumulative Deaths | Daily Cases | Daily Deaths | Cumulative Recovered | Cumulative Active Cases |
| --- | --- | --- | --- | --- | --- | --- |
| 29.03.20 | 2482 | 29 | 183 | 2 | 333 | 2105 |
| 30.03.20 | 2642 | 32 | 189 | 1 | 365 | 2223 |
| 31.03.20 | 2803 | 34 | 195 | 2 | 398 | 2342 |
| 01.04.20 | 2963 | 36 | 201 | 2 | 430 | 2460 |
| 02.04.20 | 3124 | 38 | 207 | 2 | 463 | 2578 |
| 03.04.20 | 3284 | 40 | 213 | 2 | 495 | 2697 |
| 04.04.20 | 3444 | 42 | 219 | 2 | 528 | 2815 |
| 05.04.20 | 3605 | 45 | 225 | 2 | 560 | 2934 |
| 06.04.20 | 3765 | 47 | 231 | 2 | 593 | 3052 |
| 07.04.20 | 3926 | 49 | 237 | 2 | 625 | 3170 |

**Figure 12.**
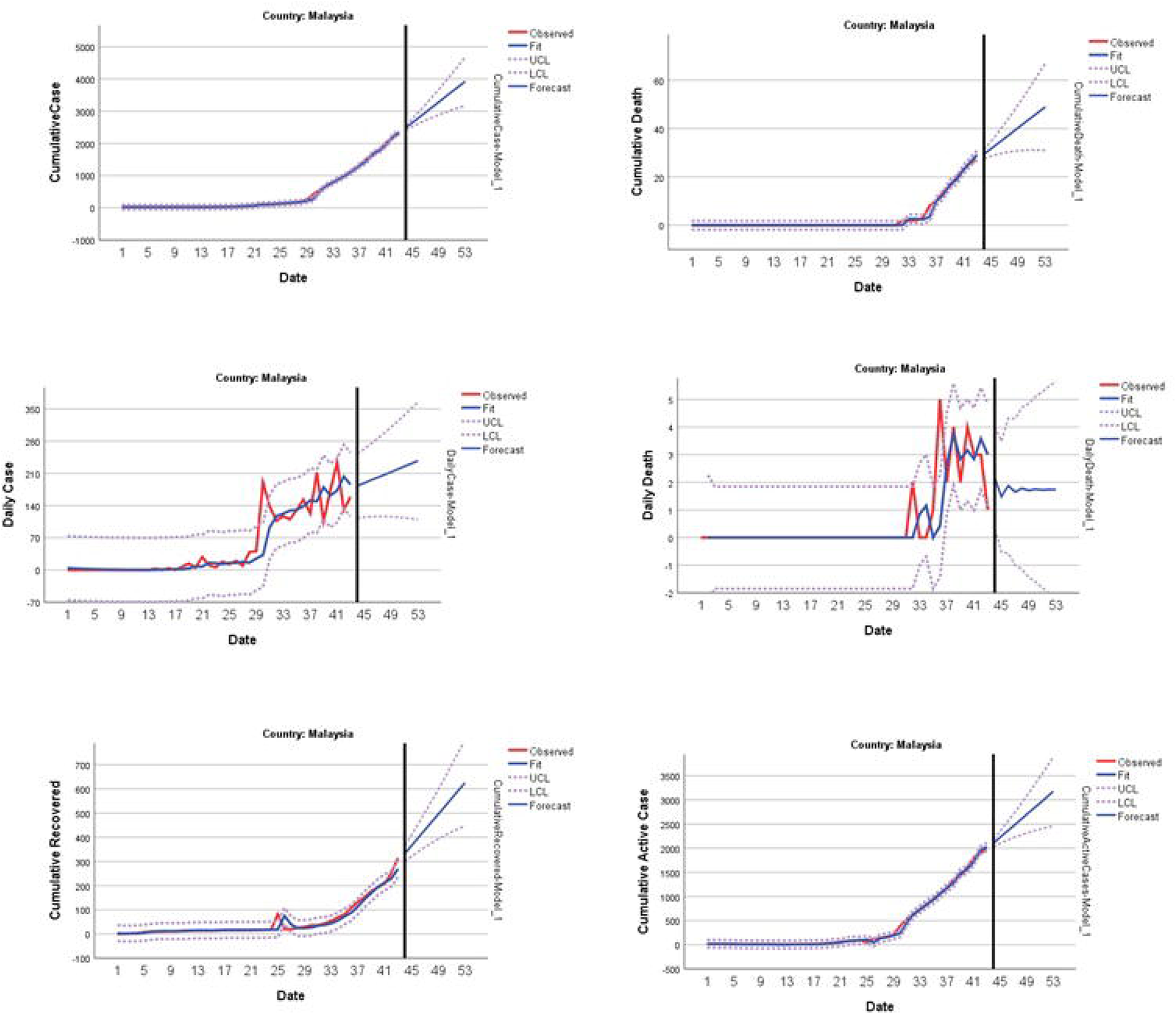
Predicting and forecasting of indicators for Malaysia

Model performance criteria obtained when Belgium data exposed to the outbreak between February 1-15 were modeled (Table 16).

**Table 16.** Times series models for Belgium that announced the first case in the period of 1-15 Feb

| Country |  | Model Type | R-squared | RMSE | Normalized BIC |
| --- | --- | --- | --- | --- | --- |
| Belgium | Cumulative Case | Brown | ,994 | 170,678 | 10,367 |
|  | Cumulative Death | Brown | ,992 | 164,562 | 10,382 |
|  | Daily Case | Brown | ,990 | 7,946 | 4,233 |
|  | Daily Death | Holt | ,983 | 32,323 | 7,039 |
|  | Cumulative Recovered | Brown | ,860 | 6,791 | 4,006 |
|  | Active Cases | Holt | ,845 | 152,513 | 10,142 |

For Belgium, it was predicted that the daily mortality prevalence would be low in the cumulative case, the daily case prevalence would be around 15% and the cumulative recovered prevalence would be around 10% in the first week of April (Table 17 and Figure 13).

**Table 17.** Forecasting for 10 days in Belgium

| Date | Cumulative Cases | Cumulative Deaths | Daily Cases | Daily Deaths | Cumulative Recovered | Cumulative Active Cases |
| --- | --- | --- | --- | --- | --- | --- |
| 29.03.20 | 10842 | 417 | 1879 | 79 | 1241 | 9035 |
| 30.03.20 | 12556 | 481 | 2113 | 90 | 1425 | 10474 |
| 31.03.20 | 14270 | 546 | 2346 | 101 | 1608 | 11912 |
| 01.04.20 | 15984 | 610 | 2579 | 112 | 1791 | 13351 |
| 02.04.20 | 17698 | 674 | 2812 | 123 | 1975 | 14789 |
| 03.04.20 | 19412 | 738 | 3045 | 135 | 2158 | 16227 |
| 04.04.20 | 21126 | 802 | 3279 | 146 | 2341 | 17666 |
| 05.04.20 | 22840 | 867 | 3512 | 157 | 2525 | 19104 |
| 06.04.20 | 24555 | 931 | 3745 | 168 | 2708 | 20542 |
| 07.04.20 | 26269 | 995 | 3978 | 179 | 2891 | 21981 |

**Figure 13.**
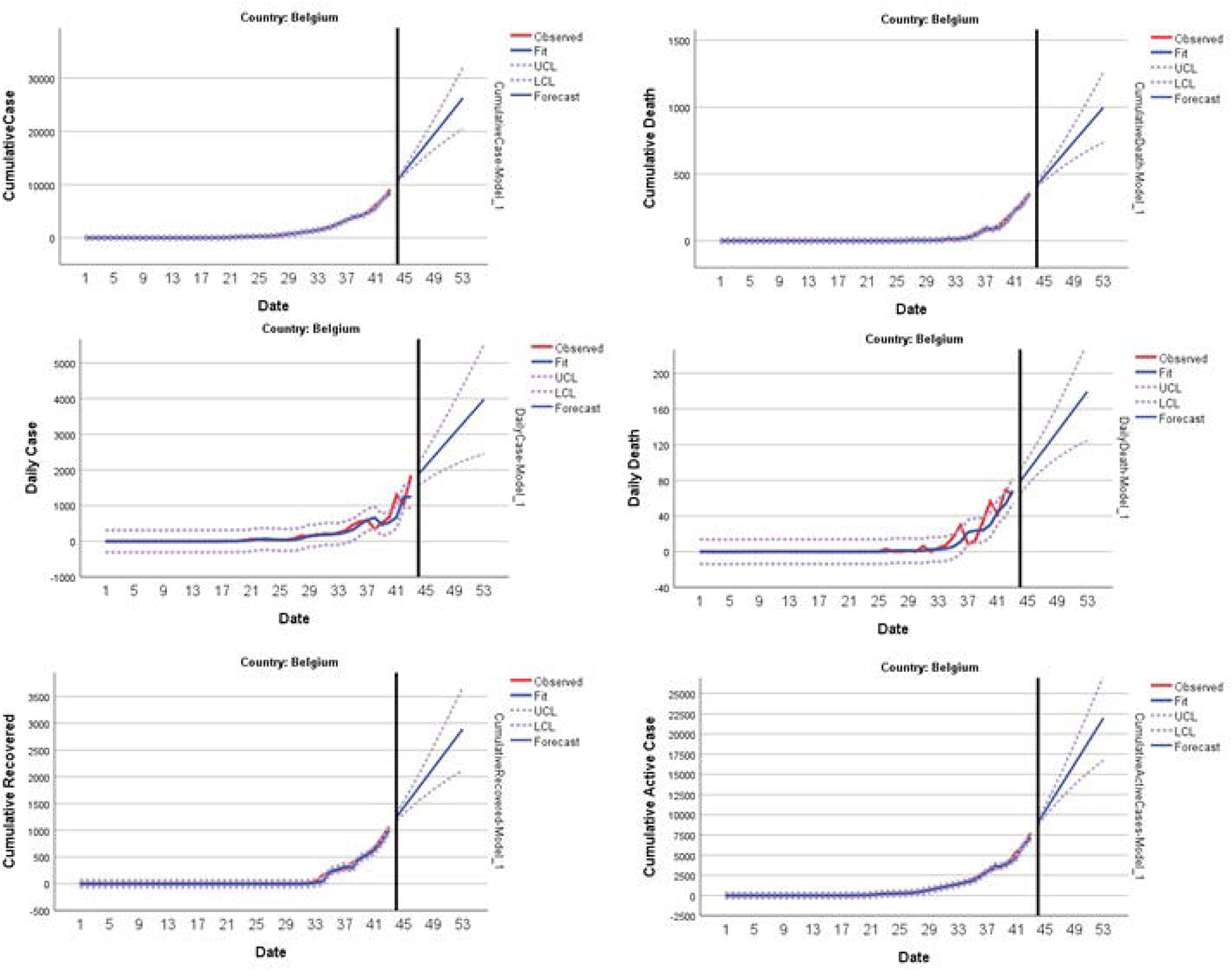
Predicting and forecasting of indicators for Belgium Iran and Israel, which were exposed to the outbreak between 16-20 February, model performance criteria obtained and the model results were given in Table 18.

**Table 18.** Times series models for Iran and Israel that announced the first case in the period of 16-20 Feb

| Country |  | Model Type | R-squared | RMSE | Normalized BIC |
| --- | --- | --- | --- | --- | --- |
| Iran | Cumulative Case | Brown | 1,000 | 9,938 | 4,687 |
|  | Cumulative Death | Brown | 1,000 | 231,627 | 10,984 |
|  | Daily Case | ARIMA(0,1,0) | ,998 | 294,052 | 11,461 |
|  | Daily Death | ARIMA(0,1,0) | ,997 | 195,074 | 10,641 |
|  | Cumulative Recovered | Brown | ,975 | 9,364 | 4,570 |
|  | Active Cases | Brown | ,921 | 219,931 | 10,882 |
| Israel | Cumulative Case | Brown | ,994 | 77,028 | 8,786 |
|  | Cumulative Death | Brown | ,993 | 76,848 | 8,781 |
|  | Daily Case | Brown | ,981 | 3,401 | 2,643 |
|  | Daily Death | ARIMA(1,0,0) | ,937 | ,770 | -,426 |
|  | Cumulative Recovered | Holt | ,797 | 72,314 | 8,660 |

When Iran data is modeled, it was predicted that the daily mortality prevalence in the cumulative case will be low in the first week of April, the daily case prevalence would be around 6% and the cumulative recovered prevalence will be around 30% (Table 19 and Figure 14).

**Table 19.** Forecasting for 10 days in Iran

| Date | Cumulative Cases | Cumulative Deaths | Daily Cases | Daily Deaths | Cumulative Recovered | Cumulative Active Cases |
| --- | --- | --- | --- | --- | --- | --- |
| 29.03.20 | 38484 | 2656 | 3157 | 143 | 12346 | 23540 |
| 30.03.20 | 41560 | 2795 | 3238 | 146 | 12990 | 25870 |
| 31.03.20 | 44636 | 2934 | 3319 | 150 | 13634 | 28200 |
| 01.04.20 | 47712 | 3073 | 3400 | 154 | 14278 | 30531 |
| 02.04.20 | 50788 | 3212 | 3481 | 157 | 14923 | 32861 |
| 03.04.20 | 53864 | 3351 | 3562 | 161 | 15567 | 35192 |
| 04.04.20 | 56940 | 3490 | 3643 | 165 | 16211 | 37522 |
| 05.04.20 | 60016 | 3629 | 3724 | 168 | 16855 | 39853 |
| 06.04.20 | 63092 | 3768 | 3805 | 172 | 17499 | 42183 |
| 07.04.20 | 66168 | 3907 | 3885 | 176 | 18143 | 44514 |

**Figure 14.**
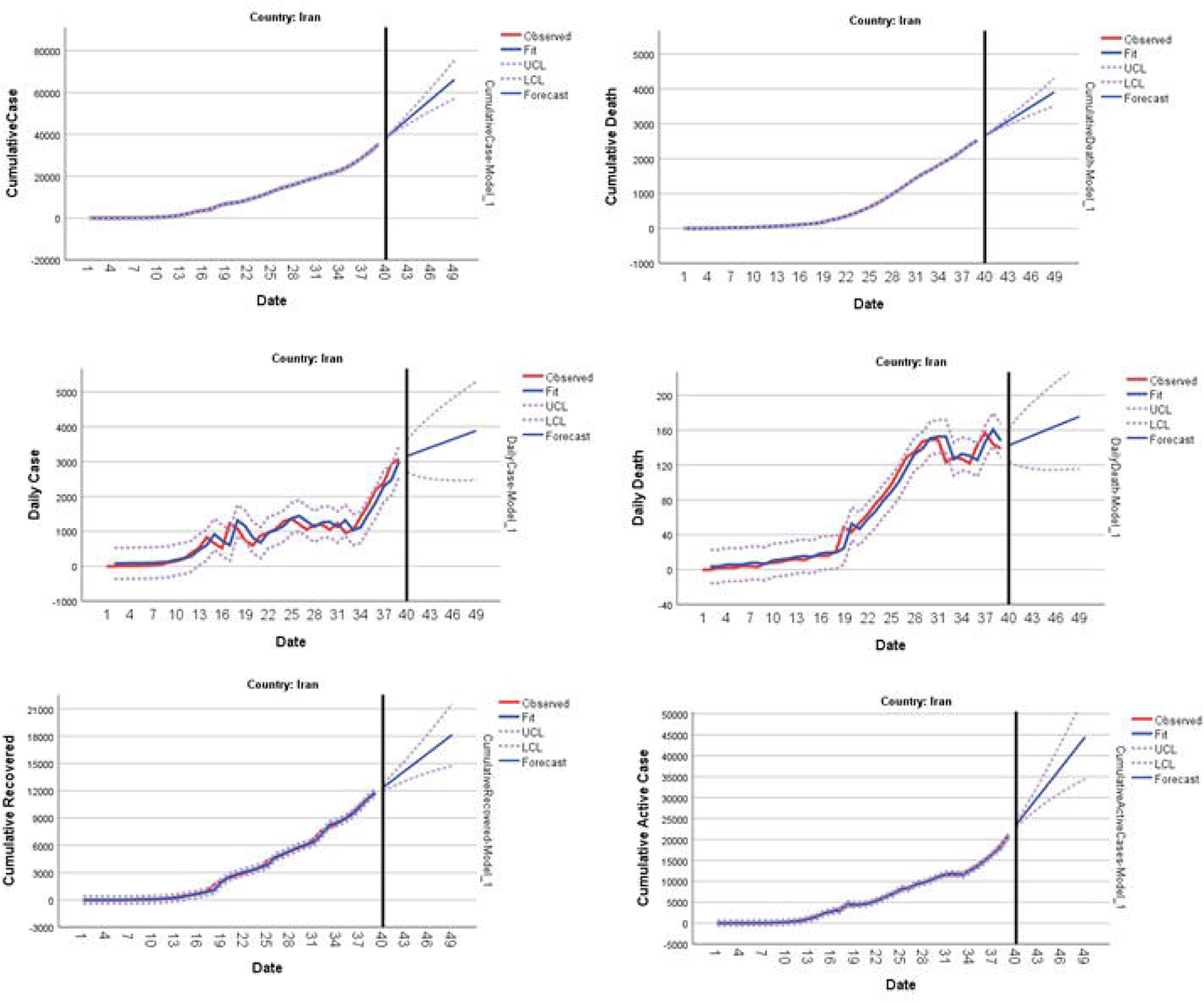
Predicting and forecasting of indicators for Iran

When Israel data was modeled, it is predicted that there would be no new daily deaths in the first week of April, the daily case prevalence in the cumulative case would be around 10% and the cumulative recovered prevalence would be around 2% (Table 20 and Figure 15).

**Table 20.** Forecasting for 10 days in Israel

| Date | Cumulative Cases | Cumulative Deaths | Daily Cases | Daily Deaths | Cumulative Recovered |
| --- | --- | --- | --- | --- | --- |
| 29.03.20 | 4130 | 14 | 527 | 0 | 96 |
| 30.03.20 | 4647 | 16 | 562 | 0 | 106 |
| 31.03.20 | 5163 | 17 | 598 | 0 | 116 |
| 01.04.20 | 5680 | 19 | 633 | 0 | 126 |
| 02.04.20 | 6197 | 21 | 668 | 0 | 136 |
| 03.04.20 | 6713 | 22 | 704 | 0 | 147 |
| 04.04.20 | 7230 | 24 | 739 | 0 | 157 |
| 05.04.20 | 7747 | 26 | 775 | 0 | 167 |
| 06.04.20 | 8263 | 27 | 810 | 0 | 177 |
| 07.04.20 | 8780 | 29 | 845 | 0 | 187 |

**Figure 15.**
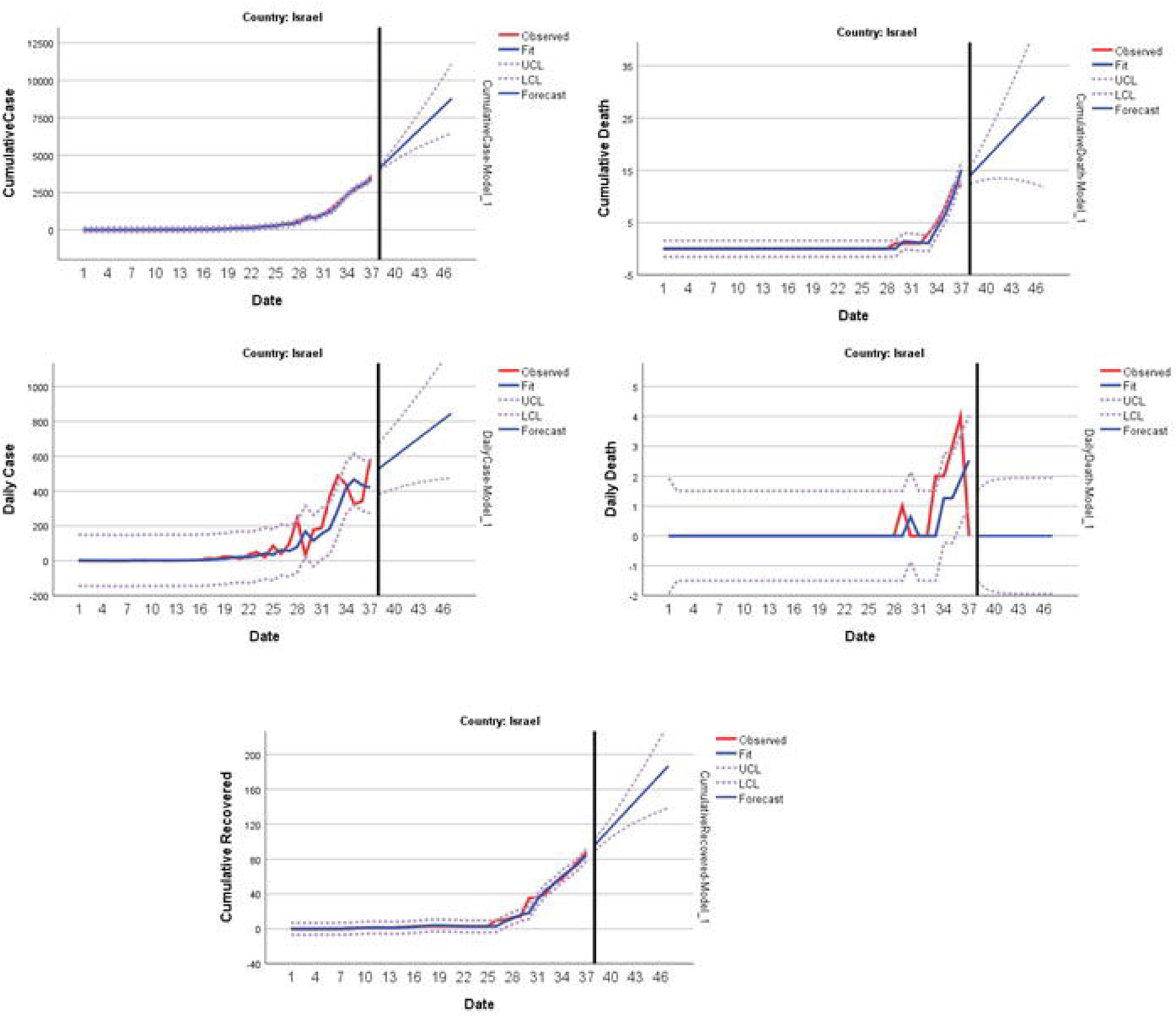
Predicting and forecasting of indicators for Israel

The model performance criteria obtained when the data of 7 countries selected among the countries exposed to the outbreak between 21-29 February were modeled are given in Table 21.

**Table 21.**
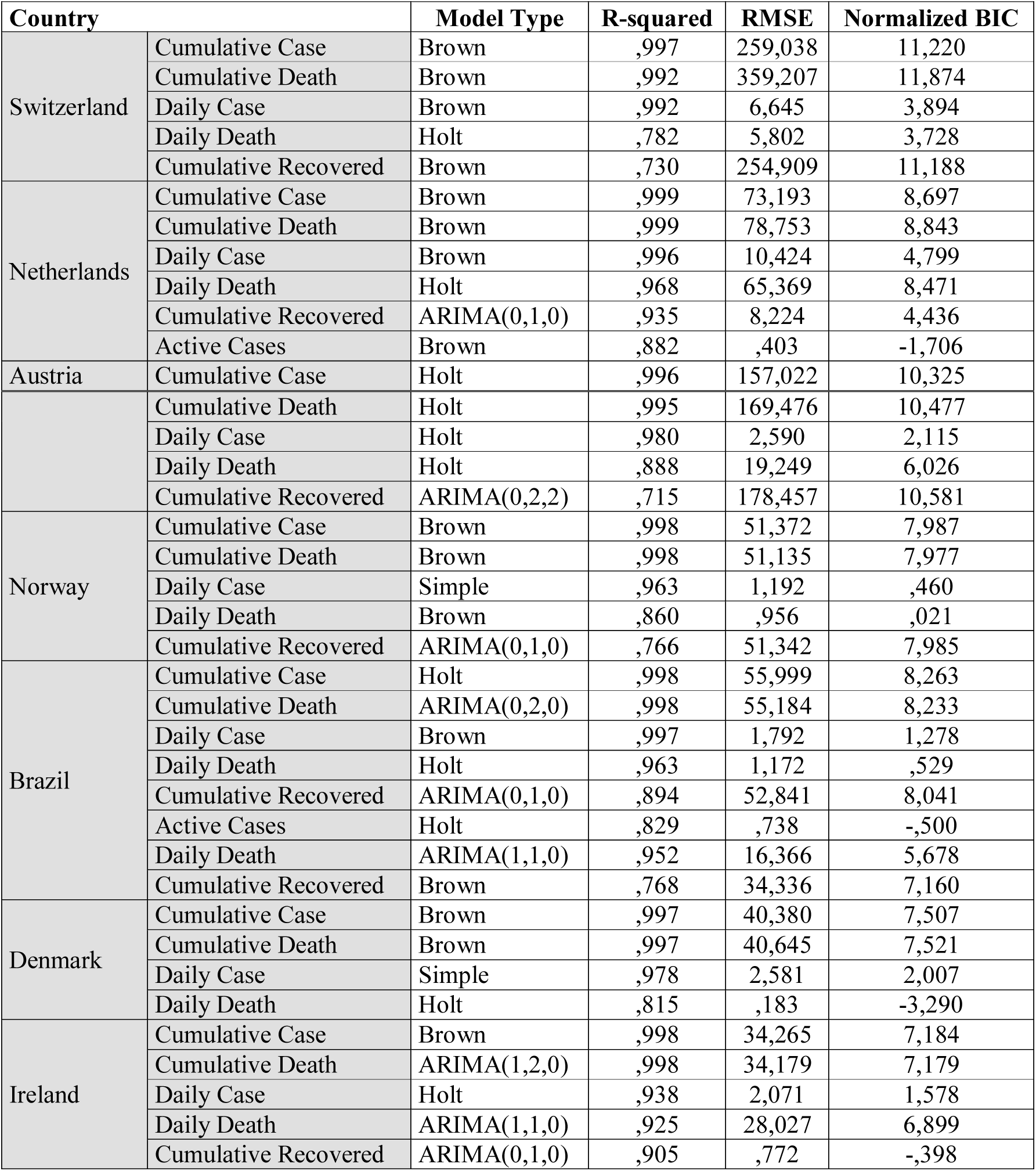
Times series models for Iran and Israel that announced the first case in the period of 21-29 February

It was predicted that the number of new daily deaths will be low in the first week of April, the daily case prevalence in the cumulative case would be around 6-10% and the cumulative recovered prevalence would be around 75% when Switzerland data was modeled (Table 22 and Figure 16).

**Table 22.** Forecasting for 10 days in Switzerland

| Date | Cumulative Cases | Cumulative Deaths | Daily Cases | Daily Deaths | Cumulative Recovered | Cumulative Active Cases |
| --- | --- | --- | --- | --- | --- | --- |
| 29.03.20 | 15177 | 299 | 1213 | 40 | 1861 | 12858 |
| 30.03.20 | 16284 | 334 | 1256 | 44 | 2202 | 13499 |
| 31.03.20 | 17392 | 368 | 1299 | 48 | 2543 | 14141 |
| 01.04.20 | 18500 | 403 | 1342 | 53 | 2884 | 14783 |
| 02.04.20 | 19608 | 438 | 1384 | 57 | 3225 | 15425 |
| 03.04.20 | 20716 | 473 | 1427 | 61 | 3566 | 16067 |
| 04.04.20 | 21824 | 507 | 1470 | 65 | 3907 | 16709 |
| 05.04.20 | 22932 | 542 | 1513 | 69 | 4248 | 17351 |
| 06.04.20 | 24039 | 577 | 1556 | 73 | 4589 | 17993 |
| 07.04.20 | 25147 | 612 | 1599 | 77 | 4930 | 18635 |

**Figure 16.**
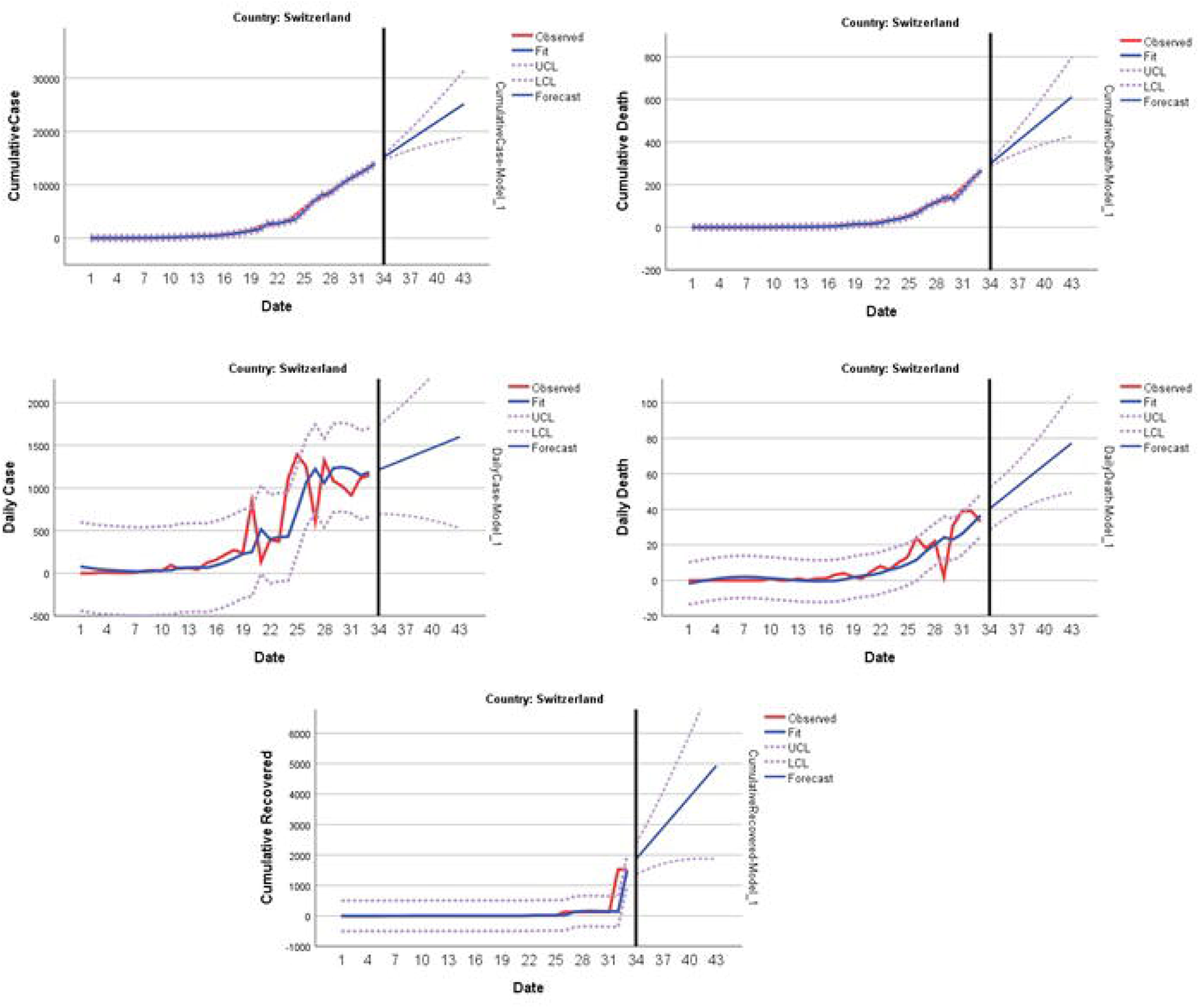
Predicting and forecasting of indicators for Switzerland

When the Netherlands results were analyzed, it was predicted that cumulative recovered and daily new deaths are very low in the first week of April, the daily case prevalence in the cumulative case would be around 8-10% and the cumulative active case prevalence would be reach around 92% (Table 23 and Figure 17).

**Table 23.** Forecasting for 10 days in Netherlands

| Date | Cumulative Cases | Cumulative Deaths | Daily Cases | Daily Deaths | Cumulative Recovered | Cumulative Active Cases |
| --- | --- | --- | --- | --- | --- | --- |
| 29.03.20 | 10921 | 734 | 1282 | 121 | 3 | 10189 |
| 30.03.20 | 12080 | 830 | 1375 | 135 | 3 | 11255 |
| 31.03.20 | 13239 | 925 | 1468 | 149 | 3 | 12321 |
| 01.04.20 | 14398 | 1020 | 1561 | 163 | 3 | 13387 |
| 02.04.20 | 15557 | 1116 | 1654 | 177 | 4 | 14453 |
| 03.04.20 | 16716 | 1211 | 1747 | 190 | 4 | 15519 |
| 04.04.20 | 17875 | 1306 | 1840 | 204 | 4 | 16585 |
| 05.04.20 | 19034 | 1402 | 1933 | 218 | 4 | 17651 |
| 06.04.20 | 20193 | 1497 | 2026 | 232 | 4 | 18717 |
| 07.04.20 | 21352 | 1592 | 2119 | 245 | 4 | 19783 |

**Figure 17.**
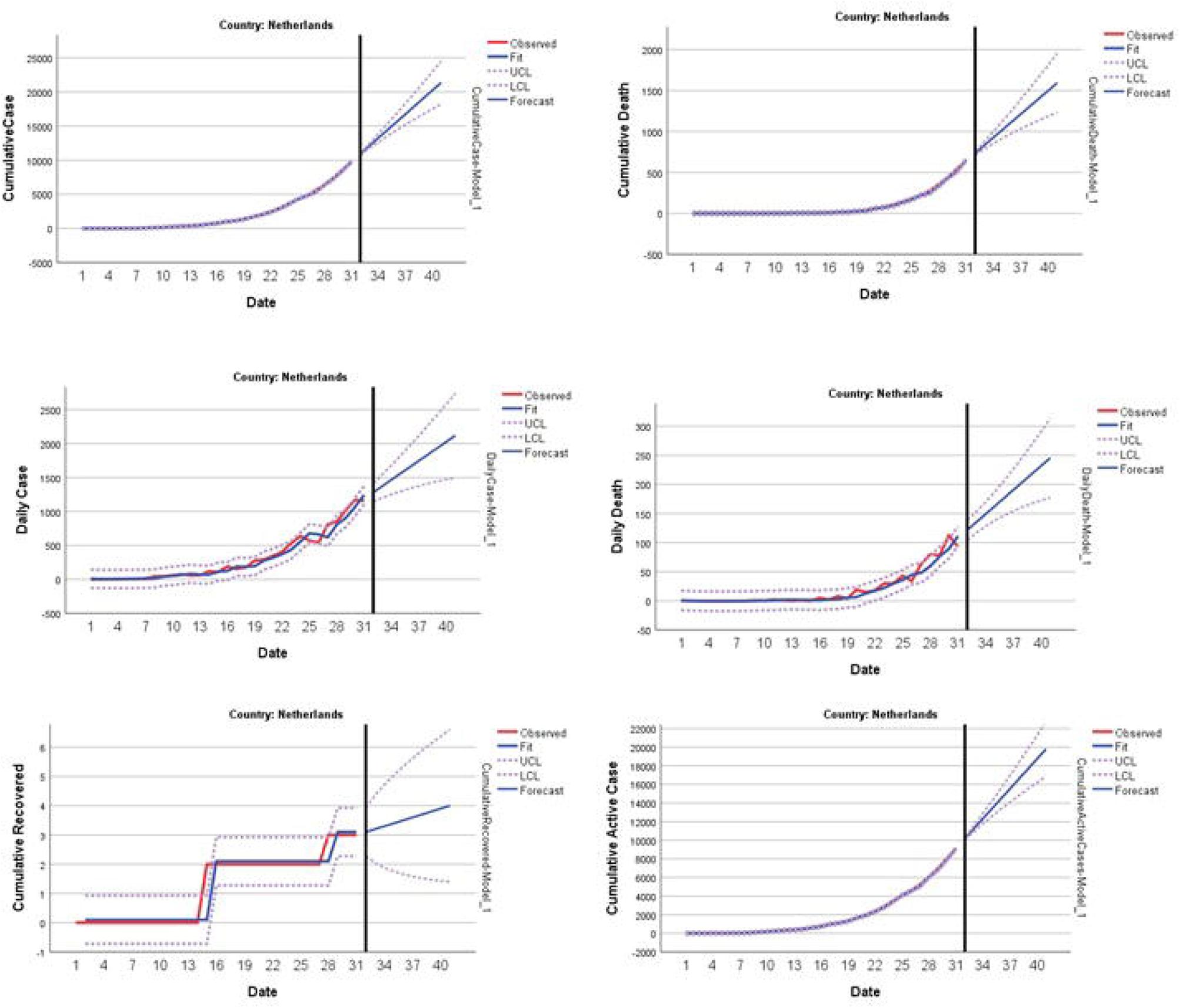
Predicting and forecasting of indicators for Netherlands

When the results of Austria data were evaluated, it was predicted that cumulative death, daily new death and cumulative recovered prevalence would be quite low in the first week of April. In addition, the cumulative recovered prevalence in the cumulative case was estimated to be around 2% (Table 24 and Figure 18).

**Table 24.** Forecasting for 10 days in Austria

| Date | Cumulative Cases | Cumulative Deaths | Daily Cases | Daily Deaths | Cumulative Recovered | Cumulative Active Cases |
| --- | --- | --- | --- | --- | --- | --- |
| 29.03.20 | 9270 | 81 | 998 | 14 | 216 | 8867 |
| 30.03.20 | 10106 | 93 | 1025 | 16 | 226 | 9601 |
| 31.03.20 | 10941 | 105 | 1052 | 18 | 236 | 10336 |
| 01.04.20 | 11777 | 117 | 1079 | 20 | 246 | 11070 |
| 02.04.20 | 12613 | 129 | 1106 | 21 | 256 | 11804 |
| 03.04.20 | 13448 | 141 | 1134 | 23 | 266 | 12539 |
| 04.04.20 | 14284 | 153 | 1161 | 25 | 275 | 13273 |
| 05.04.20 | 15120 | 165 | 1188 | 27 | 285 | 14007 |
| 06.04.20 | 15955 | 177 | 1215 | 28 | 295 | 14742 |
| 07.04.20 | 16791 | 189 | 1242 | 30 | 305 | 15476 |

**Figure 18.**
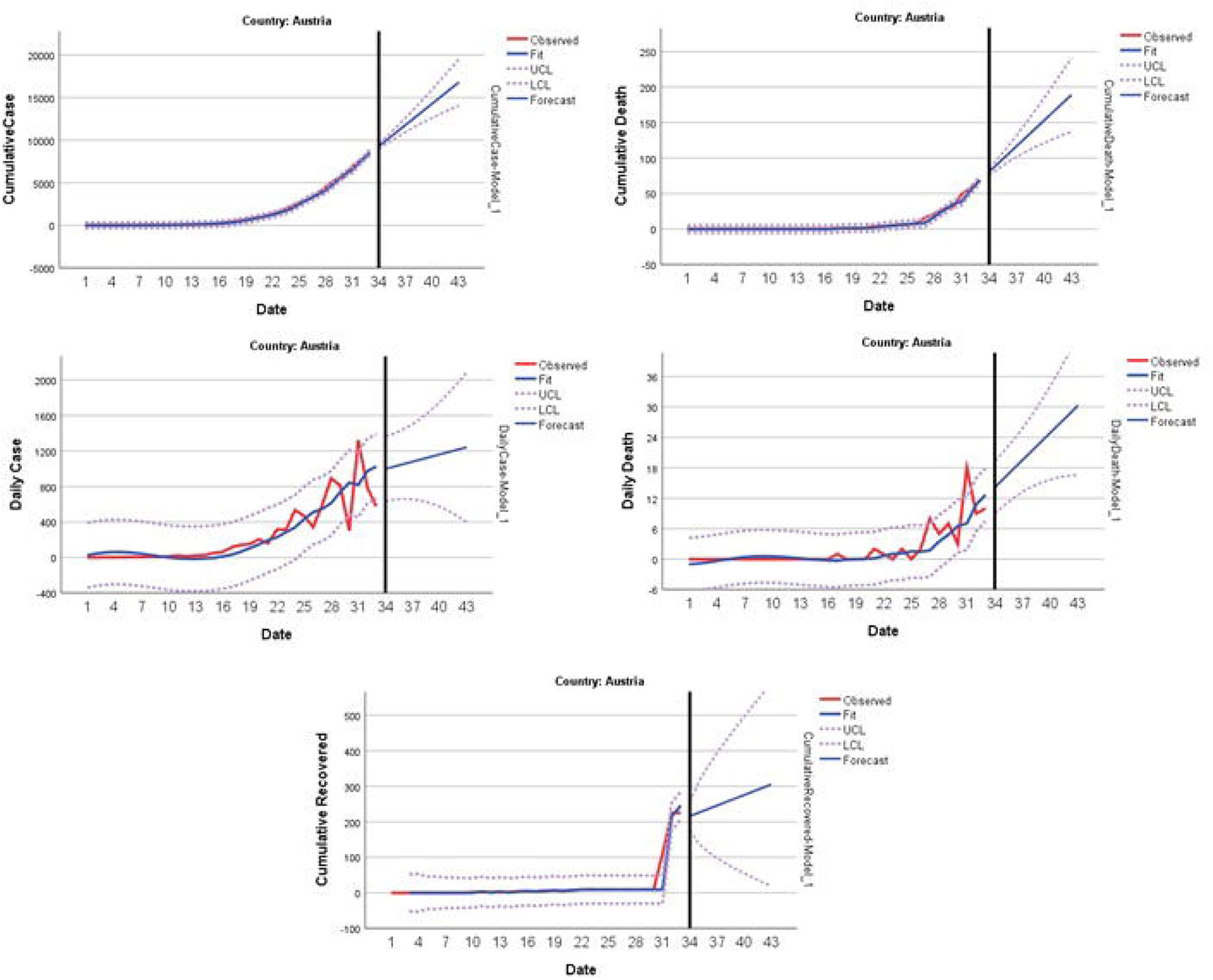
Predicting and forecasting of indicators for Austria

Cumulative death, Daily new death and cumulative recovered prevalence were predicted to be quite low in the first week of April for Norway. In addition, the prevalence of daily new cases in the cumulative case was estimated to be around 6-7% (Table 25 and Figure 19).

**Table 25.**
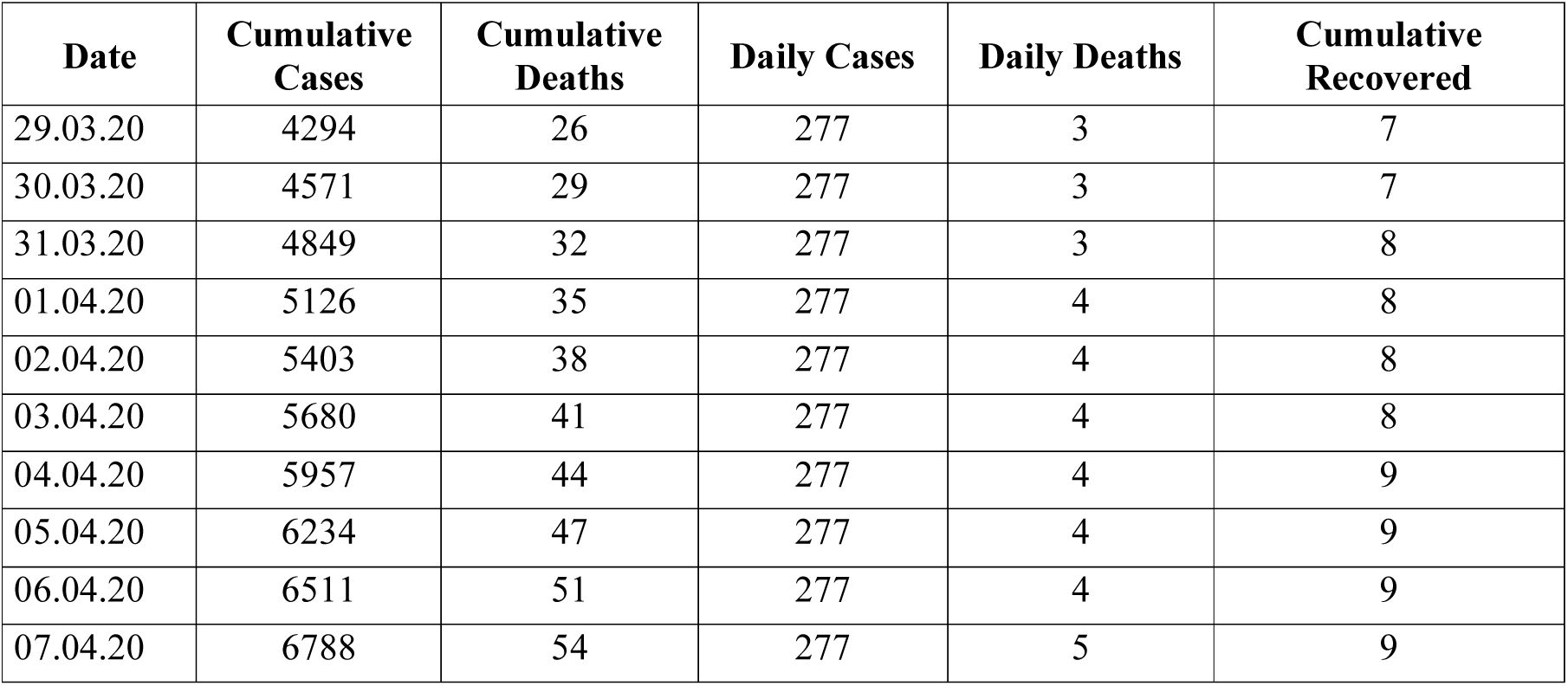
Forecasting for 10 days in Norway

**Figure 19.**
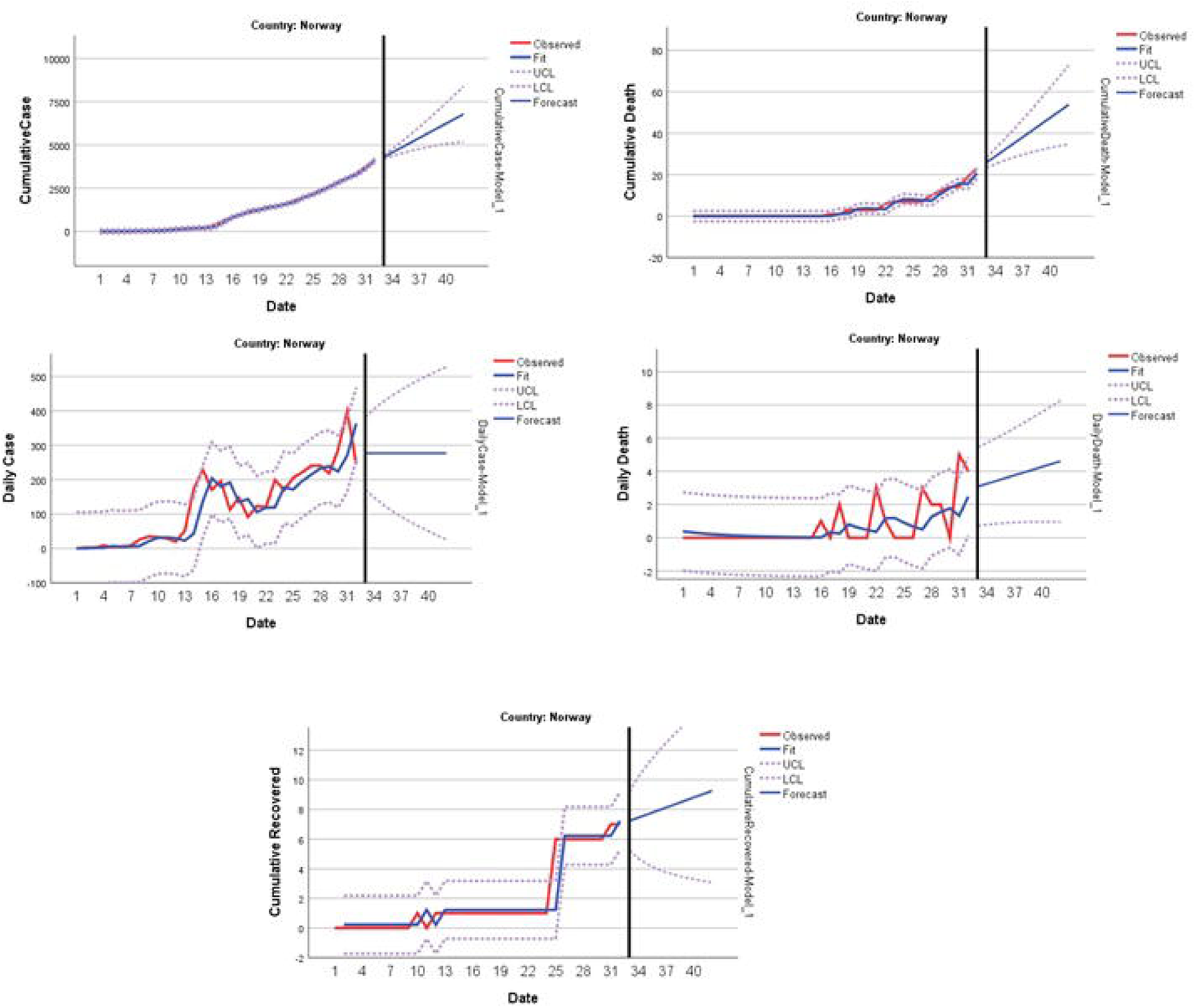
Predicting and forecasting of indicators for Norway

The prevalence of daily new death and cumulative recovered were predicted to be quite low in the first week of April for Brazil. On the other hand, the cumulative active case ratio in cumulative cases was estimated at 95% (Table 26 and Figure 20).

**Table 26.** Forecasting for 10 days in Brazil

| Date | Cumulative Cases | Cumulative Deaths | Daily Cases | Daily Deaths | Cumulative Recovered | Cumulative Active Cases |
| --- | --- | --- | --- | --- | --- | --- |
| 29.03.20 | 4371 | 137 | 502 | 23 | 6 | 4231 |
| 30.03.20 | 4860 | 160 | 532 | 25 | 6 | 4700 |
| 31.03.20 | 5349 | 184 | 562 | 28 | 7 | 5168 |
| 01.04.20 | 5838 | 209 | 592 | 30 | 7 | 5636 |
| 02.04.20 | 6326 | 235 | 622 | 33 | 7 | 6105 |
| 03.04.20 | 6815 | 261 | 652 | 35 | 7 | 6573 |
| 04.04.20 | 7304 | 288 | 682 | 38 | 7 | 7041 |
| 05.04.20 | 7793 | 316 | 712 | 40 | 8 | 7510 |
| 06.04.20 | 8282 | 344 | 742 | 43 | 8 | 7978 |
| 07.04.20 | 8771 | 373 | 772 | 45 | 8 | 8446 |

**Figure 20.**
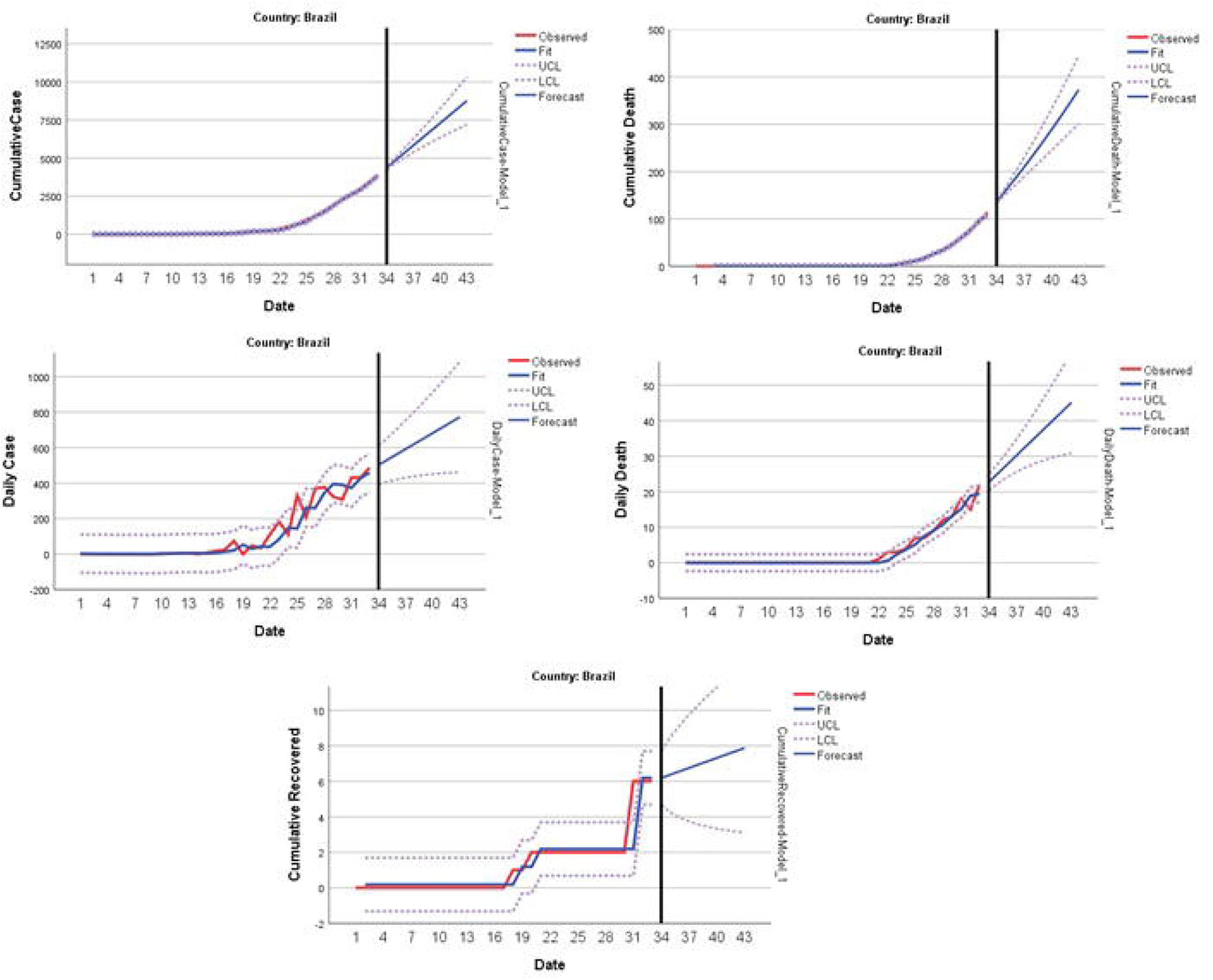
Predicting and forecasting of indicators for Brazil

According to Denmark results, the prevalence of daily new death and cumulative recovered were predicted to be quite low in the first week of April. On the other hand, the cumulative active case ratio in the cumulative case was estimated at 95% (Table 27 and Figure 21).

**Table 27.** Forecasting for 10 days in Denmark

| Date | Cumulative Cases | Cumulative Deaths | Daily Cases | Daily Deaths | Cumulative Recovered | Cumulative Active Cases |
| --- | --- | --- | --- | --- | --- | --- |
| 29.03.20 | 2356 | 75 | 155 | 12 | 1 | 2277 |
| 30.03.20 | 2511 | 86 | 155 | 13 | 1 | 2419 |
| 31.03.20 | 2666 | 97 | 155 | 15 | 1 | 2561 |
| 01.04.20 | 2821 | 108 | 155 | 16 | 1 | 2703 |
| 02.04.20 | 2976 | 119 | 155 | 18 | 1 | 2845 |
| 03.04.20 | 3131 | 130 | 155 | 19 | 1 | 2987 |
| 04.04.20 | 3286 | 141 | 155 | 21 | 1 | 3129 |
| 05.04.20 | 3441 | 152 | 155 | 23 | 1 | 3271 |
| 06.04.20 | 3596 | 163 | 155 | 24 | 1 | 3413 |
| 07.04.20 | 3751 | 173 | 155 | 26 | 1 | 3555 |

**Figure 21.**
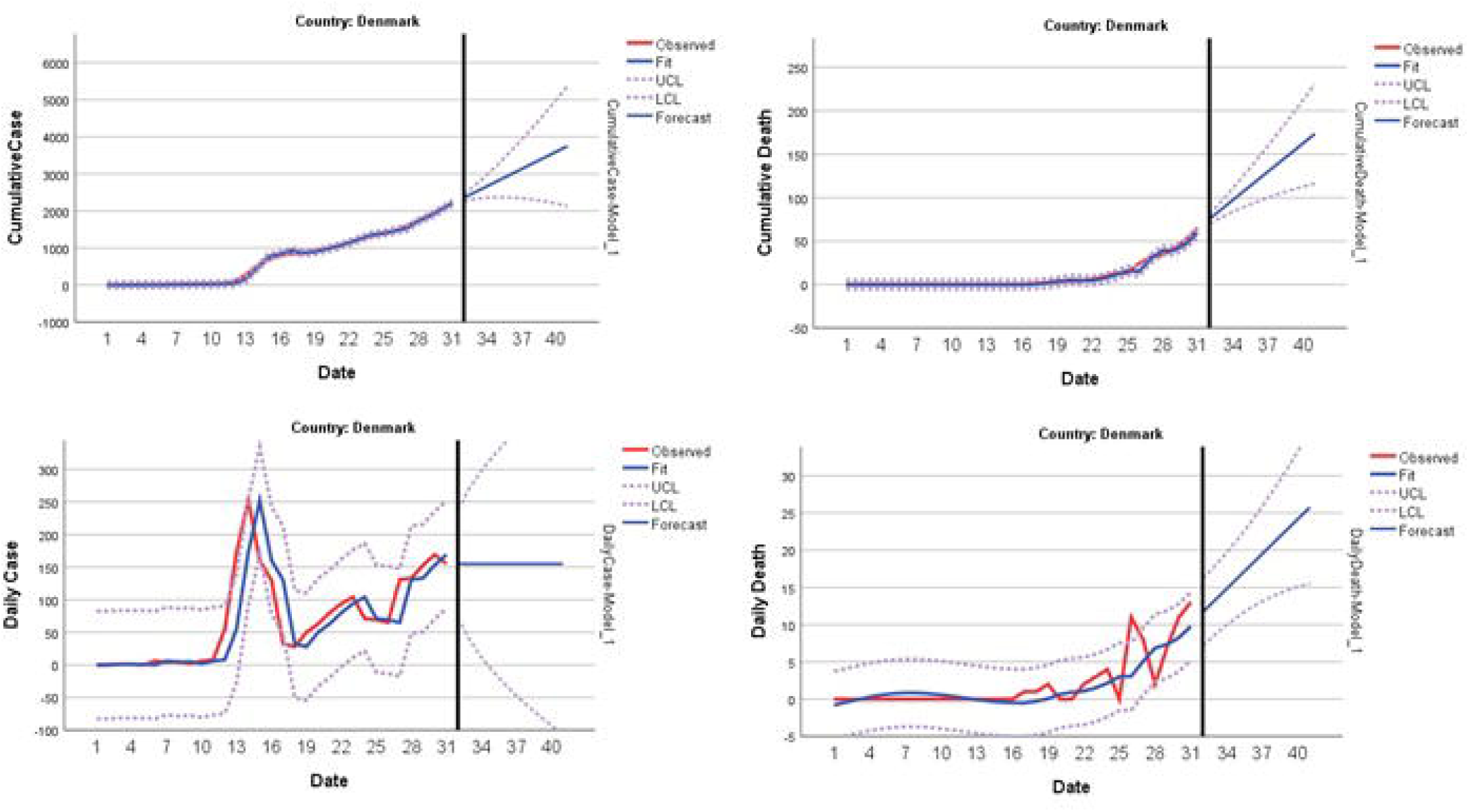
Predicting and forecasting of indicators for Denmark

It was predicted that daily new death and cumulative recovered prevalence will be quite low in the first week of April for Ireland.On the other hand, the cumulative active case prevalence in the cumulative case was estimated at 95% and the daily case prevalence was estimated at 7-10% (Table 28 and Figure 22).

**Table 28.** Forecasting for 10 days in Ireland

| Date | Cumulative Cases | Cumulative Deaths | Daily Cases | Daily Deaths | Cumulative Recovered | Cumulative Active Cases |
| --- | --- | --- | --- | --- | --- | --- |
| 29.03.20 | 2709 | 40 | 328 | 4 | 5 | 2656 |
| 30.03.20 | 3004 | 53 | 354 | 13 | 5 | 2938 |
| 31.03.20 | 3298 | 58 | 380 | 5 | 6 | 3220 |
| 01.04.20 | 3593 | 70 | 405 | 12 | 6 | 3502 |
| 02.04.20 | 3887 | 76 | 431 | 6 | 6 | 3784 |
| 03.04.20 | 4181 | 88 | 457 | 12 | 6 | 4066 |
| 04.04.20 | 4476 | 94 | 483 | 6 | 6 | 4348 |
| 05.04.20 | 4770 | 105 | 508 | 11 | 6 | 4630 |
| 06.04.20 | 5064 | 112 | 534 | 7 | 7 | 4912 |
| 07.04.20 | 5359 | 122 | 560 | 11 | 7 | 5194 |

**Figure 22.**
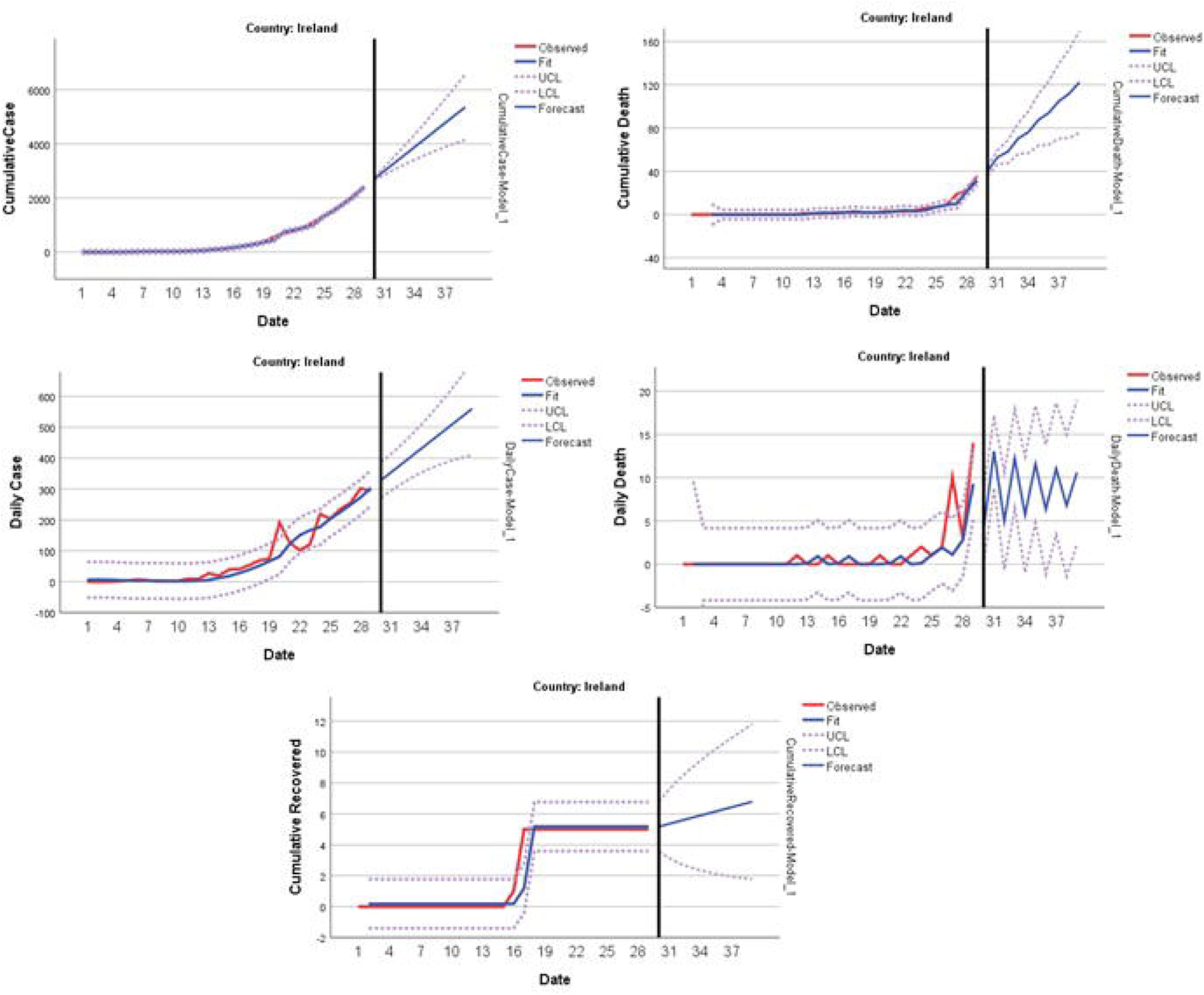
Predicting and forecasting of indicators for Ireland

Table 29 shows the model performance criteria for data from two countries selected from countries exposed to the outbreak between 1-7 March.

**Table 29.** Times series models for Iran and Israel that announced the first case in the period of 1-7 March

| Country |  | Model Type | R-squared | RMSE | Normalized BIC |
| --- | --- | --- | --- | --- | --- |
| Portugal | Cumulative Case | ARIMA(1,2,0) | ,998 | 71,667 | 8,673 |
|  | Cumulative Death | Holt | ,997 | 72,671 | 8,701 |
|  | Daily Case | Holt | ,991 | 2,550 | 2,117 |
|  | Daily Death | Holt | ,937 | 65,057 | 8,595 |
|  | Cumulative Recovered | Brown | ,925 | 1,807 | 1,427 |
|  | Active Cases | ARIMA(1,2,0) | ,901 | 4,364 | 3,069 |
| Poland | Cumulative Case | Brown | ,998 | 22,988 | 6,399 |
|  | Cumulative Death | Brown | ,998 | 23,721 | 6,461 |
|  | Daily Case | Holt | ,971 | ,957 | ,040 |
|  | Daily Death | Holt | ,950 | 15,681 | 5,762 |
|  | Cumulative Recovered | Simple | ,619 | 3,727 | 2,760 |

When Portugal’s results were analyzed, it was predicted that daily new death and cumulative recovered prevalence would be quite low in the first week of April. On the other hand, the prevalence of daily cases in the cumulative cases was at most 15% and it was predicted that this prevalence will decrease gradually, but the cumulative active cases prevalence would be 95% (Table 30 and Figure 23).

**Table 30.** Forecasting for 10 days in Portugal

| Date | Cumulative Cases | Cumulative Deaths | Daily Cases | Daily Deaths | Cumulative Recovered | Cumulative Active Cases |
| --- | --- | --- | --- | --- | --- | --- |
| 29.03.20 | 6284 | 121 | 947 | 25 | 50 | 6114 |
| 30.03.20 | 7709 | 144 | 1083 | 29 | 55 | 7503 |
| 31.03.20 | 9619 | 167 | 1219 | 34 | 60 | 9364 |
| 01.04.20 | 12249 | 190 | 1355 | 38 | 65 | 11923 |
| 02.04.20 | 16036 | 213 | 1491 | 42 | 70 | 15602 |
| 03.04.20 | 21677 | 236 | 1626 | 46 | 76 | 21071 |
| 04.04.20 | 30450 | 258 | 1762 | 50 | 81 | 29556 |
| 05.04.20 | 44673 | 281 | 1898 | 54 | 86 | 43272 |
| 06.04.20 | 68855 | 304 | 2034 | 58 | 91 | 66512 |
| 07.04.20 | 112099 | 327 | 2170 | 62 | 96 | 107899 |

**Figure 23.**
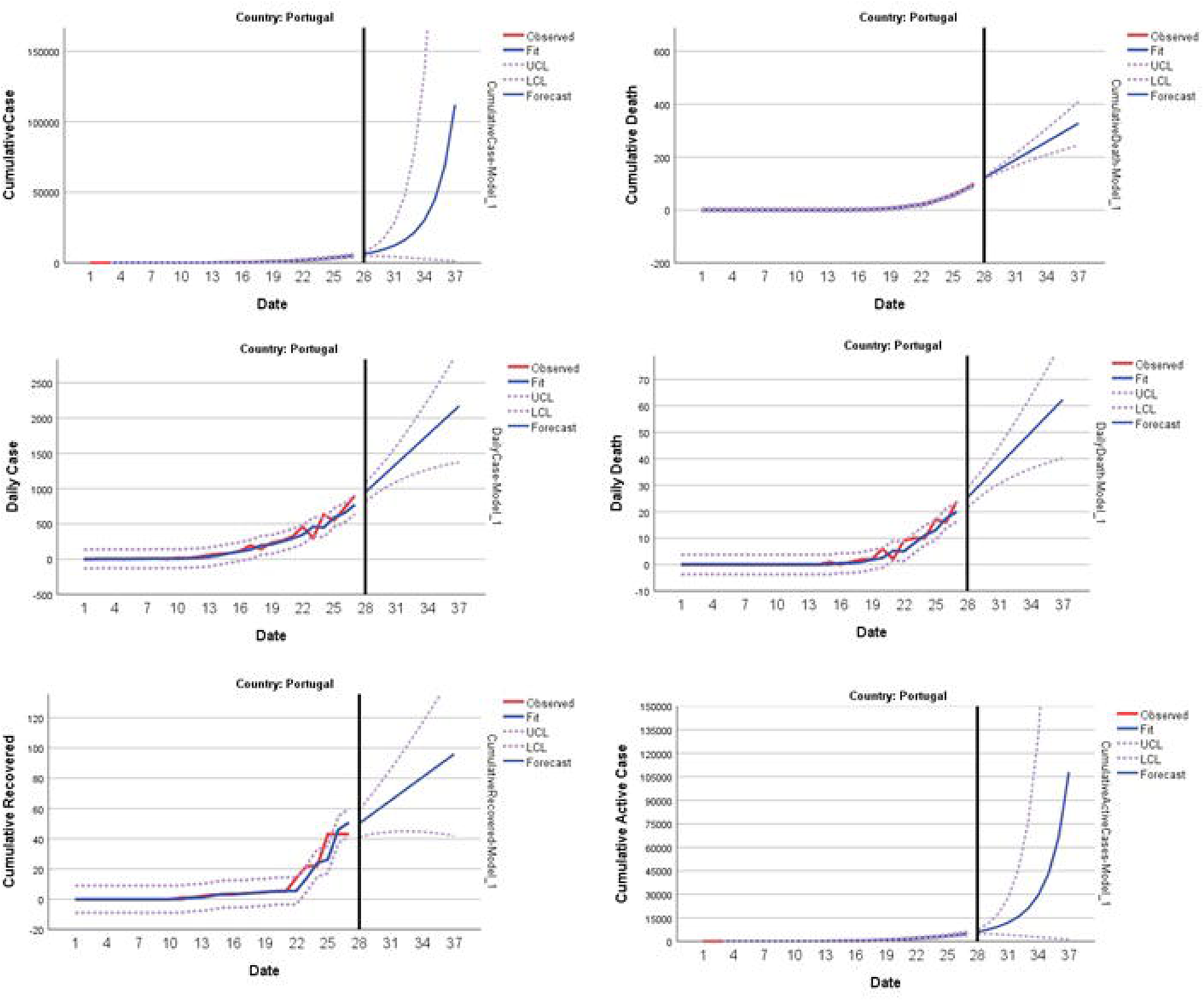
Predicting and forecasting of indicators for Portugal

It was predicted that cumulative death, daily new death and cumulative recovered prevalence would be quite low in the first week of April when Poland results were analyzed. The prevalence of daily new cases in the cumulative case was estimated at around 20% (Table 31 and Figure 24).

**Table 31.**
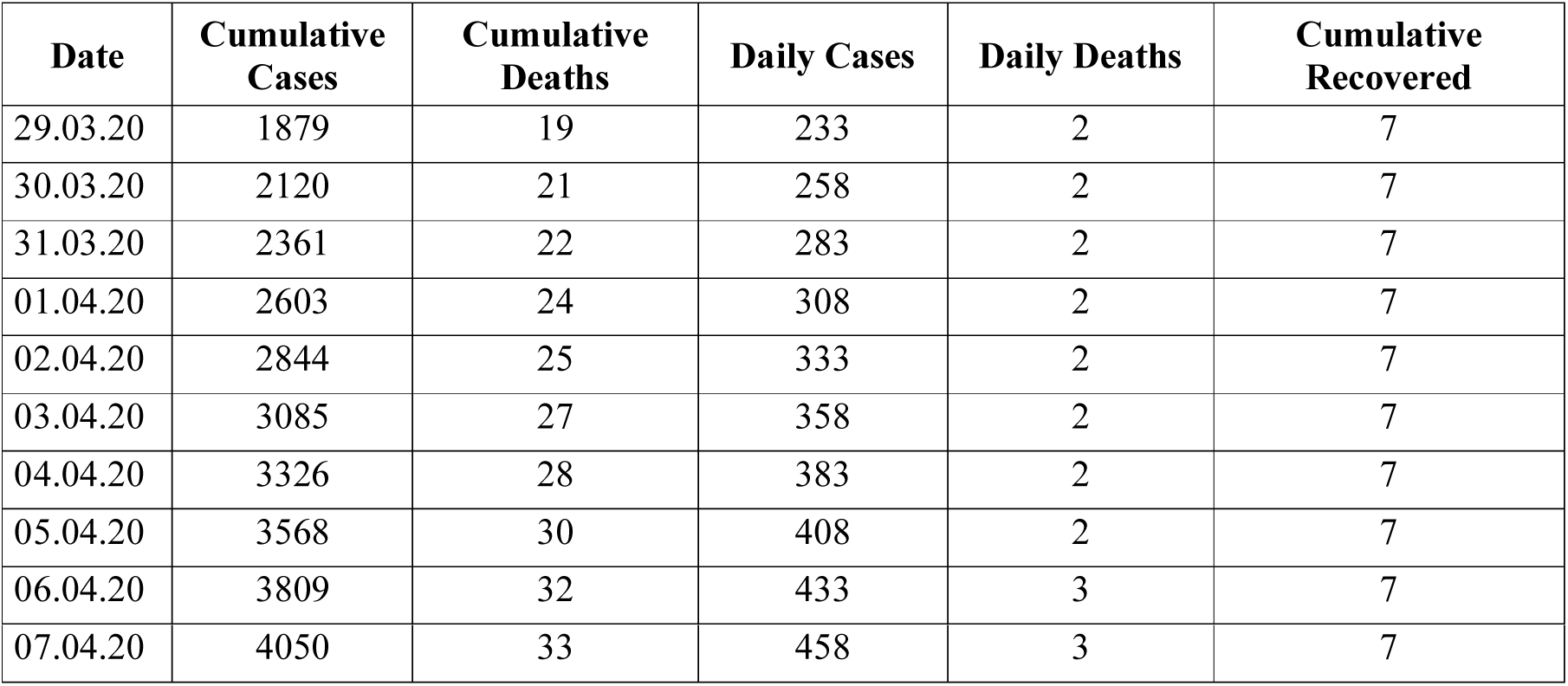
Forecasting for 10 days in Poland

| Date | Cumulative Cases | Cumulative Deaths | Daily Cases | Daily Deaths | Cumulative Recovered |
| --- | --- | --- | --- | --- | --- |
| 29.03.20 | 1879 | 19 | 233 | 2 | 7 |
| 30.03.20 | 2120 | 21 | 258 | 2 | 7 |
| 31.03.20 | 2361 | 22 | 283 | 2 | 7 |
| 01.04.20 | 2603 | 24 | 308 | 2 | 7 |
| 02.04.20 | 2844 | 25 | 333 | 2 | 7 |
| 03.04.20 | 3085 | 27 | 358 | 2 | 7 |
| 04.04.20 | 3326 | 28 | 383 | 2 | 7 |
| 05.04.20 | 3568 | 30 | 408 | 2 | 7 |
| 06.04.20 | 3809 | 32 | 433 | 3 | 7 |
| 07.04.20 | 4050 | 33 | 458 | 3 | 7 |

**Figure 24.**
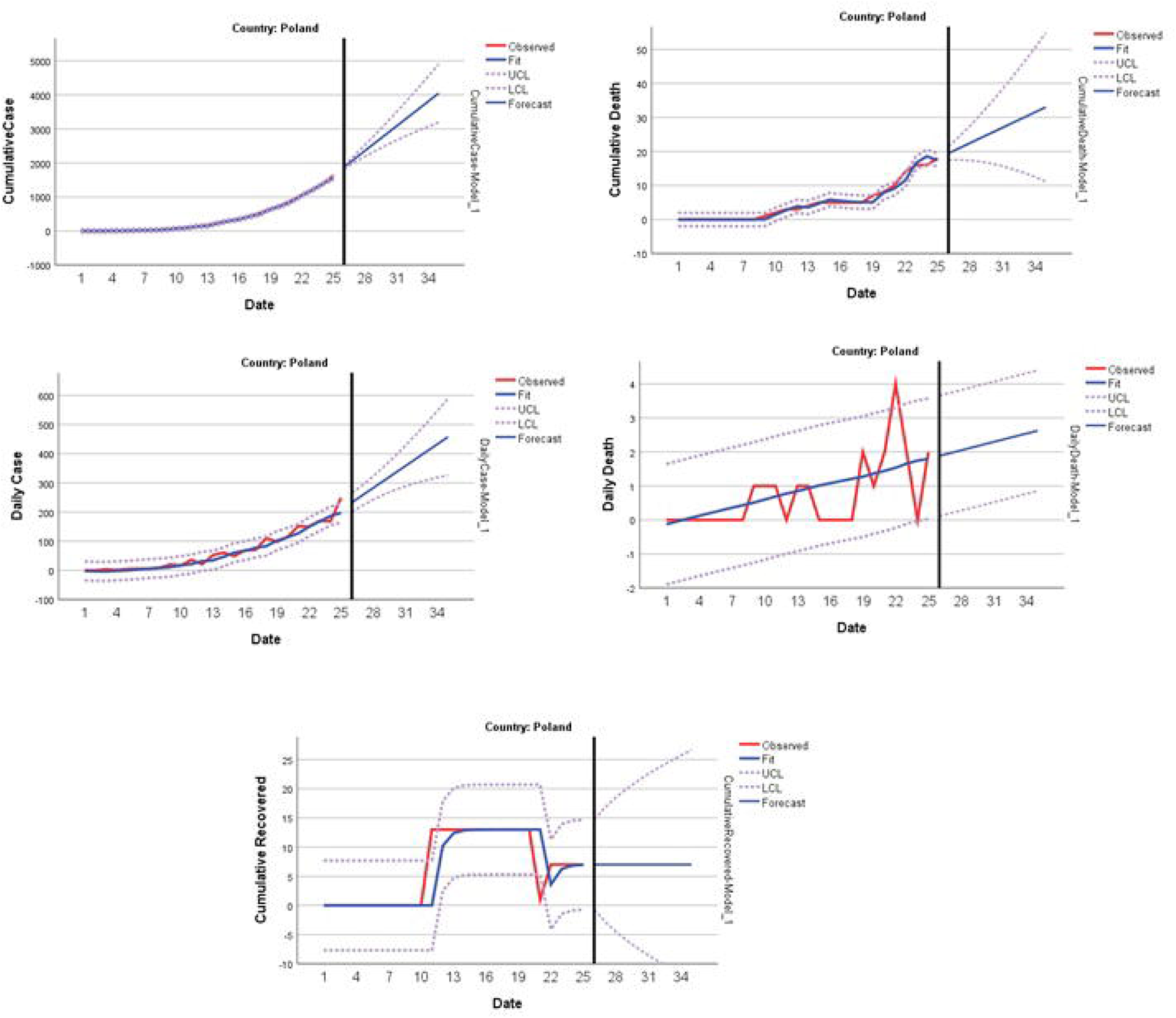
Predicting and forecasting of indicators for Poland

The model performance criteria obtained when the data of one country selected among the countries exposed to the outbreak between 8-15 March are modeled were given in Table 29.

**Table 32.** Times series models for Turkey that announced the first case in the period of 8-15 March

| Country |  | Model Type | R-squared | RMSE | Normalized BIC |
| --- | --- | --- | --- | --- | --- |
| Turkey | Total Case | ARIMA(0,1,0) | ,995 | 4,354 | 3,092 |
|  | Total Death | ARIMA(0,2,0) | ,979 | 602,322 | 13,092 |
|  | Daily Case | ARIMA(0,1,0) | ,978 | 593,394 | 13,062 |
|  | Daily Death | Brown | ,969 | 11,546 | 5,042 |
|  | Total Recovered | ARIMA(0,2,0) | ,894 | 4,092 | 2,959 |
|  | Active Cases | ARIMA(0,1,0) | ,836 | 337,764 | 11,790 |

When the results of Turkey were analyzed, it was seen that the cumulative death prevalence in the cumulative case was about 1% in the first week of April, the daily case prevalence showed a decline, the forecast was around 6% on the tenth day and the cumulative recovered prevalence was around 2% (Table 33 and Figure 25).

**Table 33.** Forecasting for 10 days in Turkey

| Date | Cumulative Cases | Cumulative Deaths | Daily Cases | Daily Deaths | Cumulative Recovered | Cumulative Active Case |
| --- | --- | --- | --- | --- | --- | --- |
| 2.04.2020 | 19741 | 343 | 2535 | 77 | 427 | 18923 |
| 3.04.2020 | 24266 | 412 | 2716 | 91 | 526 | 23196 |
| 4.04.2020 | 29120 | 484 | 2896 | 106 | 629 | 27758 |
| 5.04.2020 | 34117 | 559 | 3076 | 120 | 736 | 32426 |
| 6.04.2020 | 39022 | 637 | 3257 | 134 | 847 | 36979 |
| 7.04.2020 | 43575 | 718 | 3437 | 149 | 963 | 41166 |
| 8.04.2020 | 47505 | 802 | 3617 | 163 | 1083 | 44737 |
| 9.04.2020 | 50560 | 889 | 3798 | 178 | 1207 | 47461 |
| 10.04.2020 | 52536 | 979 | 3978 | 192 | 1336 | 49153 |
| 11.04.2020 | 53294 | 1072 | 4158 | 206 | 1469 | 49693 |

**Figure 25.**
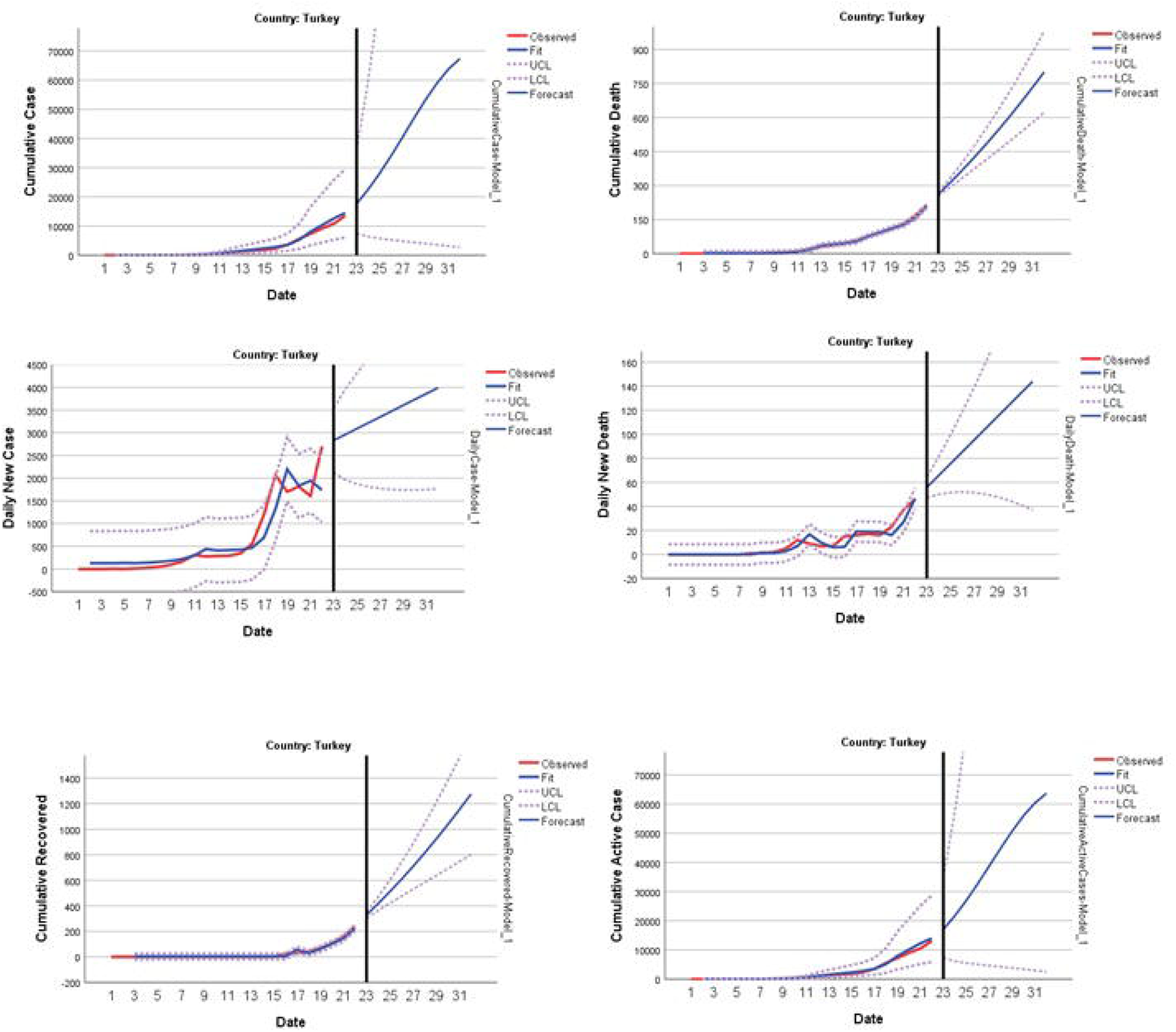
Predicting and forecasting of indicators for Turkey

In the table, on the 5th day after modeling for each country, it was calculated as the prevalence of the indicators in the cumulative cases as %. When the table was investigated, in the first week of April it was seen that the cumulative death prevalence in Italy and Spain is the highest, followed by the Netherlands, France, UK China, Denmark, Belgium, Brazil and Sweden respectively. The daily new case proportion in cumulative cases can also be considered as an indicator of an increase or decrease in the severity of the outbreak. When this indicator was evaluated, it can be said that the highest values was observed in Belgium, USA, Canada, Poland, Ireland, Netherlands, France, and Israel, which vary between 10% and 12% and the lowest proportions was observed in China and South Korea. The daily deaths proportion in the cumulative case was low in many countries and similar. But it is lowest in Netherlands. The cumulative recovered proportion in the cumulative case is the another important criteria for outbreak. This proportion was the highest in China, followed by South Korea. However, the highest value was in Switzerland (16.45%) among the ten countries exposed to the outbreak after February 21. The prevalence was found 1.75% for Turkey. Finally, the active case proportion in cumulative case was another criterion that can be used to evaluate the course of the outbreak. The predicted active case proportions are high in most of countries. This result shows that there are many people in the active disease period.

**Table 34.** The ratios of the predicted values of the indicators in the cumulative case for the first week of April

| **COVID-19**<br>**Outbreak onset**<br>**period** | **Country** | **Cum.Death**<br>**%**<br>**in**<br>**Cum. Case** | **Daily**<br>**Case %**<br>**in**<br>**Cum.**<br>**Case** | **Daily**<br>**Death**<br>**%**<br>**in**<br>**Cum.**<br>**Case** | **Cum.**<br>**Recovered**<br>**%**<br>**in**<br>**Cum. Case** | **Cum.**<br>**Active**<br>**Case %**<br>**in**<br>**Cum. Case** |
| --- | --- | --- | --- | --- | --- | --- |
| Dec 31 - Jan 15 | China | 4,07 | 0,06 | 0,01 | 95,18 | -- |
| Jan 16 - Jan 31 | Italy | 12,08 | 5,46 | 0,90 | 14,44 | 72,88 |
|  | Spain | 10,02 | 9,26 | 1,05 | 25,27 | 67,59 |
|  | South Korea | 1,73 | 1,25 | 0,07 | 63,79 | -- |
|  | Germany | 0,95 | 10,42 | 0,15 | 16,97 | 80,72 |
|  | France | 6,51 | 11,13 | 0,77 | 14,31 | 77,54 |
|  | USA | 2,89 | 14,12 | 0,48 | -- | 94,74 |
|  | UK | 6,53 | 8,61 | 1,18 | 0,30 | 92,76 |
|  | Canada | 1,14 | 11,96 | 0,14 | 7,12 | 88,13 |
|  | Sweden | 3,09 | 5,30 | 0,31 | 0,31 | -- |
|  | Australia | 0,32 | 5,70 | 0,02 | 4,66 | -- |
|  | Malaysia | 1,22 | 6,63 | 0,06 | 14,82 | 82,52 |
| Feb 1 - Feb 15 | Belgium | 3,81 | 15,89 | 0,69 | 11,16 | 83,56 |
| Feb 16 - Feb 20 | Iran | 6,32 | 6,85 | 0,31 | 29,38 | 64,70 |
|  | Israel | 0,34 | 10,78 | 0,00 | 2,19 | -- |
| Feb 21 – Feb 29 | Switzerland | 2,23 | 7,06 | 0,29 | 16,45 | 78,67 |
|  | Netherlands | 7,17 | 10,63 | 1,14 | 0,03 | 92,90 |
|  | Austria | 1,02 | 8,77 | 0,17 | 2,03 | 93,59 |
|  | Norway | 0,70 | 5,13 | 0,07 | 0,15 | -- |
|  | Brazil | 3,71 | 9,83 | 0,52 | 0,11 | 96,51 |
|  | Denmark | 4,00 | 5,21 | 0,60 | 0,03 | 95,60 |
|  | Ireland | 1,96 | 11,09 | 0,15 | 0,15 | 97,35 |
| March 1-March 7 | Portugal | 1,33 | 9,30 | 0,26 | 0,44 | 97,29 |
|  | Poland | 0,88 | 11,71 | 0,07 | 0,25 | -- |
| March 8-March 15 | Turkey | 1,18 | 8,27 | 0,23 | 1,75 | 95,60 |

\*: The calculations were made with on the 5th day after modeling for each country.

## Conclussion

The results obtained from the evaluations made with limited epidemiological data at the beginning of March or earlier have transferred some information to the present. As of the end of March, the longest 3.5 months and the shortest 1 month data are available and their evaluation is of great importance. The results of this study gives an important information for the countries struggling with the outbreak in less than one month or who have not yet been exposed to the outbreak. Also future models that can be simulated against similar risks will be obtained.

It was predicted that the outbreak in China was almost under control as a result of the models obtained with this study. The number of cases and deaths has increased rapidly in Italy and the country could not control the outbreak since the outbreak started. However, it was seen that there was a relatively increase in the proportion of the recovered. Both the rapid case increase to date and the increase seen in future forecasts show that Spain has not yet been able to control the outbreak. The new case and recovered case proportions were higher than Italy. It is seen that there has not been a significant decrease in the severity of the outbreak, although it has been 2.5 months since Italy and Spain announced the first case. South Korea, which started to fight the outbreak in the same period, was the most successful country in terms of results, followed by Germany. The results in Iran was better than Israel, which is in the same period, and France that the outbreak started 1 month ago. The results of Turkey was similar to Germany and Ireland.

The differences and advantages of this study from the modelling studies published before can be summarized as follows.

- Modeling and comparative analysis of real-time and retrospective data of 25 countries with a total number of cases exceeding 1000 among infected countries until 10 March,
- The number of data of the most countries has reached sufficient size to develop a good model
- It is also the evaluation of trend and seasonal effect in time series models.

## Data Availability

https://www.who.int/news-room/detail/08-04-2020-who-timeline---covid-19

## References

1. Jia, L., Li, K., Jiang, X. and Zhao, T. (2020). Prediction and Analysis of Coronavirus Disease 2019. Quantitative Biology. 1, 1–19.

2. Ankarali, H., Ankarali, S., Erarslan, N. (2020). COVID-19, SARS-CoV2, Infection: Current Epidemiological Analysis and Modeling of Disease. Anadolu Kliniği Tıp Bilimleri Dergisi. 25(Supplement 1), 1–22.

3. Benvenuto, D., Giovanetti, M., Vassallo, L., Angeletti and Ciccozzi, M. (2020). Application of the ARIMA model on the C0VID-2019 outbreak dataset. Data In Brief. 29, 1–4.

4. Roosa, K., Lee, Y., Luo, R., Kirpich, A., Rothenberg, R., Hyman, J.M., Yan, P. and Chowell, G. (2020). Short-term Forecasts of the COVID-19 Outbreak in Guangdong and Zhejiang, China: February 13-23, 2020. Journal of Clinical Medicine. 9, 1–9.

5. Al-qaness, M.A., Ewees, A.A., Fan, H. and El Aziz, M.M. (2020). Optimization Method for Forecasting Confirmed Cases of COVID-19 in China. Journal of Clinical Medicine. 9, 1–15.

6. Fan, C., Liu, L., Guo, W., Yang, A., Ye, C., Jilili, M., Ren, M., Xu, P., Long, H. and Wang, Y. (2020). Prediction of Outbreak Spread of the 2019 Novel Coronavirus Driven by Spring Festival Transportation in China: A Population-Based Study. International Journal of Environmental Research and Public Health. 17, 1–27.

7. Zhou, T., Liu, Q., Yang, Z., Liao, J., Yang, K., Bai, W., Lü, X. and Zhang, W. (2020). Preliminary prediction of the basic reproduction number of the Wuhan novel coronavirus 2019-nCoV. Quantitative Biology. 1,1–10.

8. Kuniya, T. (2020). Prediction of the Outbreak Peak of Coronavirus Disease in Japan, 2020. Journal of Clinical Medicine. 9, 1–7.

9. Sun, Y., Koh, V., Marimuthu, K., Ng O.T., Young, B., Vasoo, S., Chan, M., Lee, JM.V., De, P.P., Barkham, T. et al. (2020). Epidemiological and Clinical Predictors of COVID-19. Clinical Infectious Diseases. 1, 1–31.

10. Enders, W. (2004). Applied Econometric Time Series. NY: John Wiley High Education Press.

11. Franses, P.H., Dick, V.D. (2000). Non-linear Time Series Models in Empirical Finance, UK: Cambridge Univ. Press.

12. Tsay, R.S. (2010). Analysis of Financial Time Series, NY: John Wiley Press.

13. Fomby, T.B. (2008). Exponential Smoothing Models. Economics 1, 1–23.

14. Kalekar P. (2004). Time Series Forecasting Using Holt-Winters Exponential Smoothing. Information Technology. 1,1–13.

15. Hansun, S. (2016). A New Approach of Brown’s Double Exponential Smoothing Method in Time Series Analysis. Balkan Journal of Electrical & Computer Engineering. 4,75–78.

